# Comparing pre- and post-diagnosis presentations of multiple sclerosis and other inflammatory diseases in primary care: an agnostic study of French and British health records

**DOI:** 10.1101/2022.11.16.22282386

**Authors:** Octave Guinebretière, Thomas Nedelec, Laurène Gantzer, Beranger Lekens, Stanley Durrleman, Celine Louapre

## Abstract

**Importance:** The identification of a prodrome in multiple sclerosis (MS) is the key to early prevention and the targeting of new interventions.

**Objective:** To assess the associations between health conditions diagnosed in primary care and the risk of incident MS relative to other autoimmune inflammatory diseases.

**Design:** A case-control study in the UK and France was conducted from Jan 1, 1996 to March 28, 2022 in the UK and from Jan 4, 1998 to March 28, 2022 in France.

**Setting:** Data were obtained from electronic health records from the Health Improvement Network database.

**Participants:** We included all individuals with at least two years of history in the database and a recorded diagnosis of either MS, lupus or Crohn’s disease. Three controls matched for sex, age at index date and year at index date were randomly assigned to each individual with a diagnosis of MS.

**Main outcome measures:** We agnostically tested the associations between 113 different diagnoses and multiple sclerosis diagnosis during the five years before or the five years after the diagnosis of MS. Unadjusted odds ratios (ORs) and 95% CIs were estimated, and *p* values were corrected for multiple comparisons. We also stratified for sex, age at diagnosis, and year of diagnosis. A logistic regression analysis (adjusted for sex and age at diagnosis) was performed to compare MS patients with lupus and Crohn’s disease patients.

**Results:** The study population consisted of patients with MS (UK: 15,808; and France: 4,366), Crohn’s disease (UK: 20,872; and France: 9,605) or lupus (UK: 5,296; and France: 2,041). We identified twelve health conditions as significantly positively associated with the risk of MS. After considering health conditions suggestive of demyelinating events as the first diagnosis of MS, five health conditions remained significantly associated with MS: depression (UK OR 1.22 [95%CI 1.11-1.34]), sexual dysfunction (1.47 [1.11-1.95]), constipation (1.5 [1.27-1.78]), cystitis (1.21 [1.05-1.39]), and urinary tract infections (1.38 [1.18-1.61]). However, none of these conditions was selectively associated with MS in comparisons with both lupus and Crohn’s disease. During the five years after MS diagnosis, all five health conditions identified here were still associated with MS.

**Conclusion and relevance:** The identified symptoms may be considered to be not only prodromal, but also early-stage symptoms, albeit not specific to MS.

**Key points:** *Question:* What prediagnostic manifestations of multiples sclerosis (MS) occur in primary care settings and how do they differ from those of other autoimmune diseases?

*Results:* In this agnostic study of 15,808 MS patients from the UK and 4,366 MS patients from France, we identified five health conditions as positively associated with the risk of MS when recorded in the five years preceding MS diagnosis. We show that most of these health conditions were also present in the early presentations of lupus and Crohn’s disease.

*Meaning:* Our findings suggest that the prodromal phase of MS is largely similar to the prediagnostic manifestations of other autoimmune diseases.

## Introduction

There is a growing body of evidence to suggest that patients with multiple sclerosis (MS) have asymptomatic brain lesions ^1,2^ and/or serum or CSF biomarkers before the onset of typical MS symptoms.^3,4^ However, it remains a matter of debate whether this preclinical phase triggers subtle, non-specific symptoms. These health conditions would not be sufficiently pronounced to warrant diagnosis, but they might indicate the start of the neurodegenerative process, as in other neurodegenerative diseases.^5,6^ The recognition of a prodromal period would help to gain knowledge on the consequences of the appearance and accumulation of MS lesions. Additionally, in population with a high risk of developing MS due to familial history or in the case of radiologically isolated syndrome, the identification of prodromal symptoms could lead to stratify patients at higher risk of developing MS who would benefit from early therapeutic intervention.

Previous studies have reported pre-diagnostic features, such as depression, fatigue, sleep disorders or urinary infections, associated with MS and occurring up to 10 years before diagnosis.^7–9^ However, most studies on the MS prodrome have focused on either hypothesis-driven searches for non-specific symptoms,^8,9^ or major disease categories.^7^ It is also important to compare the early presentation of MS with other inflammatory diseases, to determine which of the pre-diagnosis features are specific to MS. There is also a lack of analysis of differences in the clinical presentation of the MS prodrome by age, sex,^10^ and year of diagnosis.^11^

We set up and analyzed two nested case-control studies based on data from two large primary care databases in the UK and France, extracted from The Health Improvement Network (THIN) database. We sought to confirm the expected associations between pre-diagnosis features and the subsequent diagnosis of MS, and to compare presentations before and after diagnosis. We also aimed to compare MS patient cohorts with control cohorts of patients with other autoimmune diseases — lupus and Crohn’s disease — to determine whether the features identified were specific to MS or common to several autoimmune diseases.

## Methods

We used The Health Improvement Network (THIN) database,^13^ a large standardized European database of fully anonymized and non-extrapolated electronic medical records collected from physicians by the company GERS SAS^13^ and coded according to the International Classification of Diseases, 10th revision (ICD10). The THIN database complies with all current European data protection laws (General Data Protection Regulation) and adheres to the Observational Medical Outcomes Partnership model.^13^ The French data were collected from a pool of 2500 general practitioners (GP) ^14^, and were representative of the French population in terms of age, sex, and area of residence. The UK data were collected from 400 general practices covering about 6% of the UK population, selected for the THIN quality data-recording scheme with Vision practice management software.^15^ Several studies have already reported that the electronically coded diagnoses in this database are representative of the UK general practice population in terms of demographics and type of consultation.^13^ For each patient, the diagnosis corresponding to each visit, the prescriptions made by the GP, and all other diagnoses associated with these prescriptions were available. Data for education level and social status were not provided, to preserve anonymity.

We considered data recorded from Jan 1, 1996 to March 28, 2022 in the UK and from Jan 1, 1996 to March 28, 2022 in France. In both countries, we identified all the patients with an ICD-10 code for MS (G35). The earliest date at which the G35 code was noted was taken as the index date. Age at MS onset was defined as age at the index date. For each country, we randomly selected three times as many controls as cases, with no history of MS diagnosis and a similar distribution of age at index date, sex and year at index date to the cases. A symptom was considered to be present if either the corresponding ICD-10 code or the prescription of a symptom-specific drug was recorded (eTable 1 and eTable 2). Only individuals in the case and control cohorts for whom at least two years of data before the index date were available were included, as shown in the flowchart in eFigure 1, 2.

For patients in the MS cohort with a demyelinating event recorded in the five years preceding MS diagnosis, we used the earliest recorded demyelinating event as the index date (primary analysis). We identified demyelinating events in the data on the basis of the ICD 10 codes listed in eTable 3. We also changed the index date for each matched control accordingly. We defined past exposure to the health conditions considered, based on the ICD-10 codes provided directly in both the French and UK databases. We used the first three characters of the code, defining disease category. In this exploratory approach, we assessed the association between MS and each of the health conditions defined by ICD-10 codes with a relative frequency of more than 0.5% in both MS and control cohorts and in both countries.

We also ran a sensitivity analysis, in which we shifted the index date further back, to the earliest diagnosis suggestive of MS relapse. For each country, we identified diagnoses suggestive of MS relapse as the ICD 10 codes associated with MS in the primary analysis that could be interpreted as symptoms of a relapse of MS or the first symptoms of progressive MS. We used the ICD-10 codes listed in eTable 4. We applied the same shift in index date to each matched control. We investigated the influence of sex and age before and after MS diagnosis. In the post-diagnosis analysis, individuals for whom at least two years of data after the index date were available were selected. We split the data into three cohorts of equal size, using the 33^rd^ and 66^th^ percentiles for age at diagnosis as cutoff points, for pre- and post-diagnosis analyses of the influence of age at diagnosis. New diagnostic criteria redefining approaches to MS in light of improvements in our understanding of the disease and technological breakthroughs emerged in 2010 ^17^ and 2017.^18^ These criteria have been shown to facilitate earlier diagnosis and to decrease diagnostic error and delays.^19,20^ We investigated the effect of these changes in the criteria for MS diagnosis on our findings, by assessing the effect of year of diagnosis on pre and post-diagnosis analyses, using the significant associations identified in the sensitivity analysis.

### Statistical analysis

We performed a logistic regression analysis with correction for the duration of follow-up to analyze the association between each health condition corresponding to an ICD-10 code and MS diagnosis. We quantified the association by calculating odds ratios (ORs) and 95% CIs for exposure to a particular health condition within five years of the index date. *P* values were corrected for multiple comparisons with the Bonferroni method, with an alpha risk of 5%.

For the comparison between MS and other autoimmune diseases, we included patients with lupus or Crohn’s disease as additional control cohorts. We performed a logistic regression analysis controlled for age at index date and sex. We studied post-diagnosis presentation, using a Cox regression model with MS as the exposure of interest, considering the given health condition as the outcome and the time to the event or the time to last visit or data extraction as the time variable. In investigations of temporal associations, we also used the significant findings from the comparison of the MS cohort to patients without MS to chart the single-year prevalence of the prescription of symptom-specific drugs for each of the 15 years before and after MS diagnosis.

## Results

### Demographic information

We included 4,366 and 15,763 individuals diagnosed with MS, 12,531 and 38,098 individuals without a diagnosis of MS, 2,041 and 5,296 patients diagnosed with lupus, and 9,605 and 20,872 patients diagnosed with Crohn’s disease, from France and the UK, respectively, in the analysis. All these individuals were selected from the THIN database. The lupus cohort contained slightly older patients and included more women than the MS cohort, whereas the Crohn’s disease cohort contained younger patients, with fewer women than the MS cohort. Median age at MS diagnosis was 46 years (37–57 years) in the UK and 44 years (34–54 years) in France, with an age at first MS symptoms of 44 years (34 – 54 years) in the UK and 42 years (32 – 52 years) in France. The age at first MS symptom decreased to 39 years (30 – 50 years) in the UK and 40 years (30 – 51 years) in France for patients diagnosed after 2017. More women than men were diagnosed with MS in both countries. More follow-up data were available for the UK sample on average, with 9 person-years of data available before the index date in the full analytical cohort versus only 6 person-years on average for the French sample. The median number of visits per year was also higher in the UK than in France. Demographic information for patients and controls is summarized in Table 1.

**Table 1.** Characteristics of patients with MS, Crohn’s disease, or lupus, and controls. The data are shown as percentages or medians (IQR).

|  | Size, n | Sex<br>(female) | Age at diagnosis | Age at first<br>symptom of MS | Person-years of<br>data available<br>before diagnosis | Number of<br>visits per year<br>before<br>diagnosis |
| --- | --- | --- | --- | --- | --- | --- |
| ***UK*** |  |  |  |  |  |  |
| **MS** |  |  |  |  |  |  |
| Whole cohort | 15808 | 70.5% | 46 (37 - 57) | 44 (34 - 54) | 9 (5 - 13) | 6 (2 - 7) |
| Year of MS diagnosis |  |  |  |  |  |  |
| <2010 | 10108 | 70.9% | 48 (38 - 58) | 46 (37 - 56) | 7 (5 - 10) | 6 (2 - 7) |
| [2010,2017] | 4319 | 69.9% | 44 (35-54) | 41 (31 - 51) | 13 (8 - 18) | 6 (2 - 7) |
| >2017 | 1381 | 69.1% | 44 (34 - 55) | 39 (30 - 50) | 17 (9 - 23) | 5 (2 - 7) |
| Lupus | 5296 | 88.2% | 48 (36 - 61) |  | 9 (5 - 14) | 8 (3 - 10) |
| Crohn's disease | 20872 | 54.5% | 42 (27 - 59) |  | 7 (4 - 12) | 6 (2 - 8) |
| Controls | 42226 | 70.2% | 46 (36 - 56) |  | 7 (4 - 12) | 5 (2 - 6) |
| ***France*** |  |  |  |  |  |  |
| **MS** |  |  |  |  |  |  |
| Whole cohort | 4366 | 70.2% | 44 (34 - 54) | 42 (32 - 52) | 6 (3 - 10) | 4 (2 - 6) |
| Year of MS diagnosis |  |  |  |  |  |  |
| <2010 | 1890 | 69.8% | 45 (35 - 55) | 44 (34 - 55) | 4 (3 - 6) | 5 (2 - 7) |
| [2010,2017] | 1708 | 70.4% | 43 (34 - 52) | 41 (32 - 50) | 8 (4 - 11) | 4 (2 - 5) |
| >2017 | 768 | 71.0% | 43 (32 - 54) | 40 (30 - 51) | 10 (5 - 15) | 3 (1 - 5) |
| Lupus | 2041 | 81.5% | 47 (36 - 60) |  | 7 (4 - 10) | 5 (2 - 7) |
| Crohn's disease | 9605 | 55.9% | 40 (27 - 55) |  | 7 (4 - 11) | 4 (1 - 5) |
| Controls | 12564 | 70.2% | 43 (33 - 53) |  | 4 (3 - 7) | 3 (1 - 4) |

### Primary and sensitivity analyses

Overall, 113 health conditions were considered in the French and UK databases (eTable 1). We identified 12 health conditions as significantly positively associated with a diagnosis of MS when recorded in the five years before MS diagnosis, in both countries (eTable 6). Only one of these conditions was a neurologic disorder: epilepsy and recurrent seizures. Two psychiatric disorders, depression and sexual dysfunction (not caused by organic disorder or disease), were more frequently recorded in patients with MS than in controls. The other associated health conditions were related to disorders of the vestibular system, functional intestinal disorders (principally constipation), limb pain, cystitis, urinary tract infection, skin paresthesia, dizziness and giddiness, headache, and malaise and fatigue. Skin paresthesia, dizziness and giddiness, and disorders of vestibular function may be interpreted as MS symptoms that were not recognized as such by the GP. Only major depressive disorder, sexual dysfunction, constipation, cystitis, and urinary tract infection remained significantly more frequent in patients with MS than in controls in the sensitivity analysis (Figure 1, eTable 6). Overall, 91.2% of the patients diagnosed with functional intestinal disorders in the UK and 92.4% of those diagnosed with such disorders in France had constipation. All health conditions considered to be associated with MS diagnosis in the pre-diagnosis analysis were also associated with MS diagnosis in the post-diagnosis analysis (Table 2).

**Table 2.** ORs for all health problems individually associated with MS in the sensitivity analysis during the five years before and five years after the occurrence of the first symptoms. We used the whole sensitivity cohort, sex-based cohorts, age-at-onset cohorts, and year-at-diagnosis cohorts. Results are shown for the UK cohort. Results from the French cohort is shown in eTable 7. .* cannot be calculated because too few presentations were recorded. * for ORs associated with corrected p-values between 0.05 and 0.0001. ** for ORs associated with corrected p-values below 0.0001.

|  | Whole cohort | Sex |  | Age at onset |  |  | Year of diagnosis |  |  |
| --- | --- | --- | --- | --- | --- | --- | --- | --- | --- |
|  |  | Women | Men | <40 | [40,53] | >53 | <2010 | [2010,2017] | >2017 |
| Before diagnosis |  |  |  |  |  |  |  |  |  |
| F32 : Major depressive disorder, single episode | 1.22 (1.11 - 1.34)** | 1.17 (1.08 - 1.27)** | 1.39 (1.20 - 1.60)** | 1.18 (1.04 - 1.33)* | 1.27 (1.14 - 1.42)** | 1.19 (1.06 - 1.35)* | 1.26 (1.16 - 1.38)** | 1.21 (1.07 - 1.37)** | 1.15 (0.93 - 1.42) |
| F52 : Sexual dysfunction | 1.47 (1.11 - 1.95)** | 1.01 (0.57 - 1.79) | 1.62 (1.29 - 2.04)** | 1.85 (1.11 - 3.06)* | 1.77 (1.25 - 2.52)** | 1.2 (0.88 - 1.64) | 1.47 (1.11 - 1.95)* | 1.51 (1.06 - 2.15)* | 1.51 (0.72 - 3.19) |
| K59 : Functional intestinal disorders | 1.5 (1.27 - 1.78)** | 1.39 (1.21 - 1.61)** | 1.93 (1.48 - 2.52)** | 1.17 (0.91 - 1.49) | 1.57 (1.24 - 2.00)** | 1.74 (1.44 - 2.10)** | 1.98 (1.62 - 2.42)** | 1.48 (1.21 - 1.80)** | 1.23 (0.90 - 1.69) |
| N30 : Cystitis | 1.21 (1.05 - 1.39)* | 1.12 (1.00 - 1.26)* | 2.43 (1.69 - 3.48)** | 1.07 (0.89 - 1.30) | 1.1 (0.90 - 1.33) | 1.47 (1.23 - 1.75)** | 1.35 (1.17 - 1.57)** | 1.15 (0.96 - 1.39) | 1.13 (0.84 - 1.51) |
| N39 : Urinary tract infection, site not specified | 1.38 (1.18 - 1.61)** | 1.26 (1.11 - 1.43)** | 2.39 (1.75 - 3.26)** | 1.12 (0.90 - 1.39) | 1.24 (1.00 - 1.54)* | 1.73 (1.45 - 2.08)** | 1.47 (1.28 - 1.69)** | 1.05 (0.82 - 1.34) | 1.32 (0.85 - 2.05) |
| After diagnosis |  |  |  |  |  |  |  |  |  |
| F32 : Major depressive disorder, single episode | 1.47 (1.40 - 1.54)** | 1.37 (1.30 - 1.45)** | 1.83 (1.65 - 2.02)** | 1.39 (1.28 - 1.52)** | 1.4 (1.29 - 1.51)** | 1.51 (1.38 - 1.65)** | 1.42 (1.34 - 1.50)** | 1.4 (1.26 - 1.55)** | 1.29 (0.95 - 1.75) |
| F52 : Sexual dysfunction | 2.51 (2.21 - 2.87)** | 1.36 (0.93 - 2.00) | 2.71 (2.36 - 3.11)** | 3.95 (2.96 - 5.28)** | 2.7 (2.21 - 3.31)** | 1.73 (1.39 - 2.16)** | 2.32 (2.02 - 2.67)** | 3.43 (2.43 - 4.85)** | * |
| K59 : Functional intestinal disorders | 2.72 (2.56 - 2.90)** | 2.58 (2.40 - 2.77)** | 3.1 (2.74 - 3.51)** | 3.0 (2.62 - 3.45)** | 3.33 (2.99 - 3.72)** | 2.3 (2.09 - 2.52)** | 2.69 (2.51 - 2.89)** | 2.37 (2.06 - 2.73)** | 2.37 (1.45 - 3.87)** |
| N30 : Cystitis | 2.13 (2.00 - 2.26)** | 1.93 (1.80 - 2.05)** | 4.3 (3.60 - 5.14)** | 2.07 (1.83 - 2.34)** | 2.17 (1.96 - 2.40)** | 2.19 (1.99 - 2.41)** | 2.14 (2.00 - 2.30)** | 1.77 (1.55 - 2.03)** | 1.82 (1.19 - 2.80)* |
| N39 : Urinary tract infection, site not specified | 2.72 (2.52 - 2.93)** | 2.46 (2.27 - 2.66)** | 4.47 (3.69 - 5.40)** | 2.42 (2.08 - 2.81)** | 3.05 (2.68 - 3.46)** | 2.8 (2.50 - 3.14)** | 2.71 (2.50 - 2.93)** | 2.55 (2.06 - 3.15)** | 3.25 (1.23 - 8.60)* |

**Figure 1.**
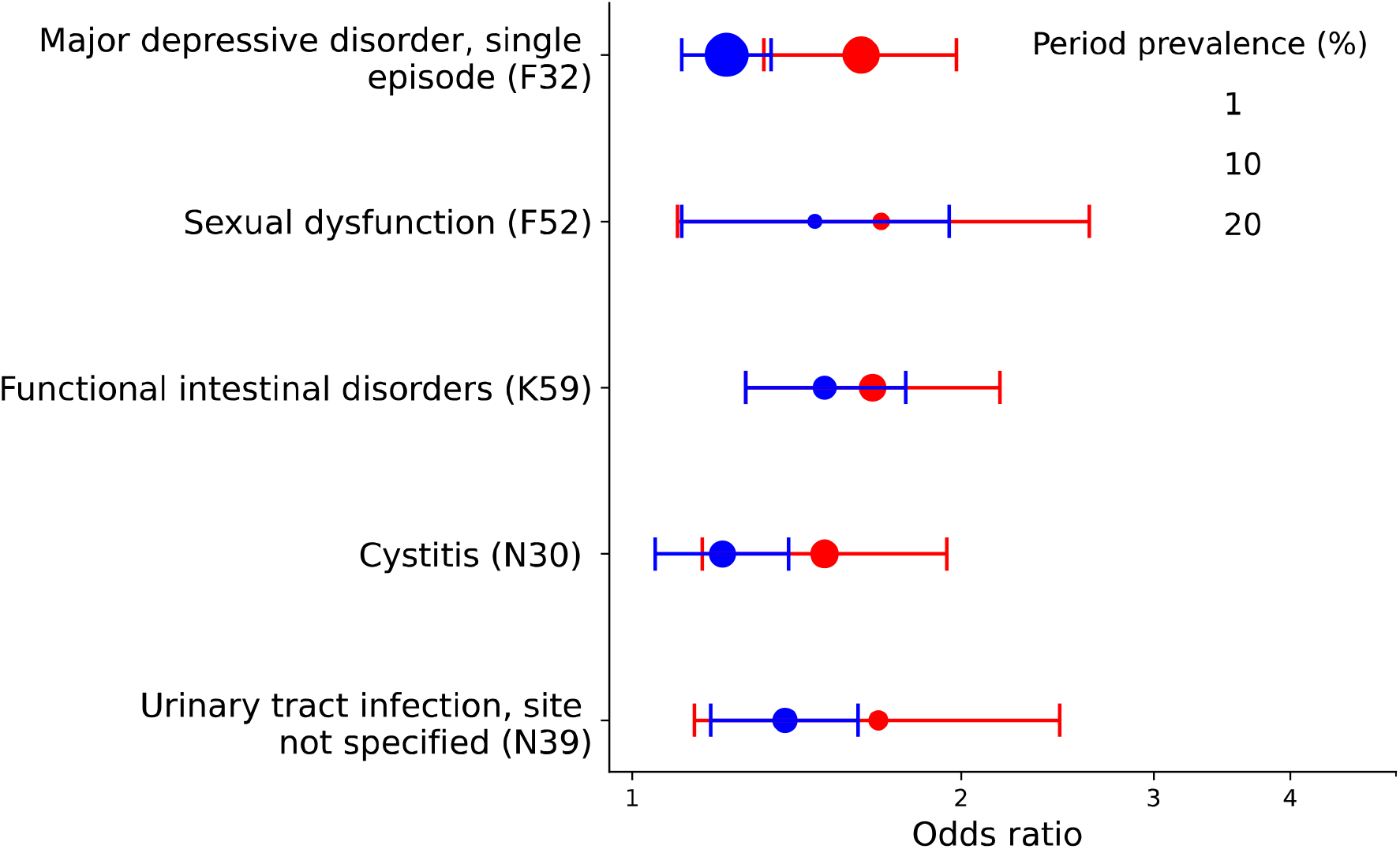
ORs for significant associations in the sensitivity analysis for five years before the index date. Only associations significant in both the French and UK cohorts are shown, with red and blue dots, respectively. The size of the dot is sproportional to the number of affected people. Bars correspond to 95% CIs after correction for multiple comparison.

### Comparison with other autoimmune diseases

Depression, sexual dysfunction, and urinary tract infection were found to be more frequent among people subsequently diagnosed with MS than in Crohn’s disease patients, in both France and the UK (Figure 2). However, no health condition was found to be significantly associated with MS in comparisons with lupus patients, in either country.

**Figure 2.**
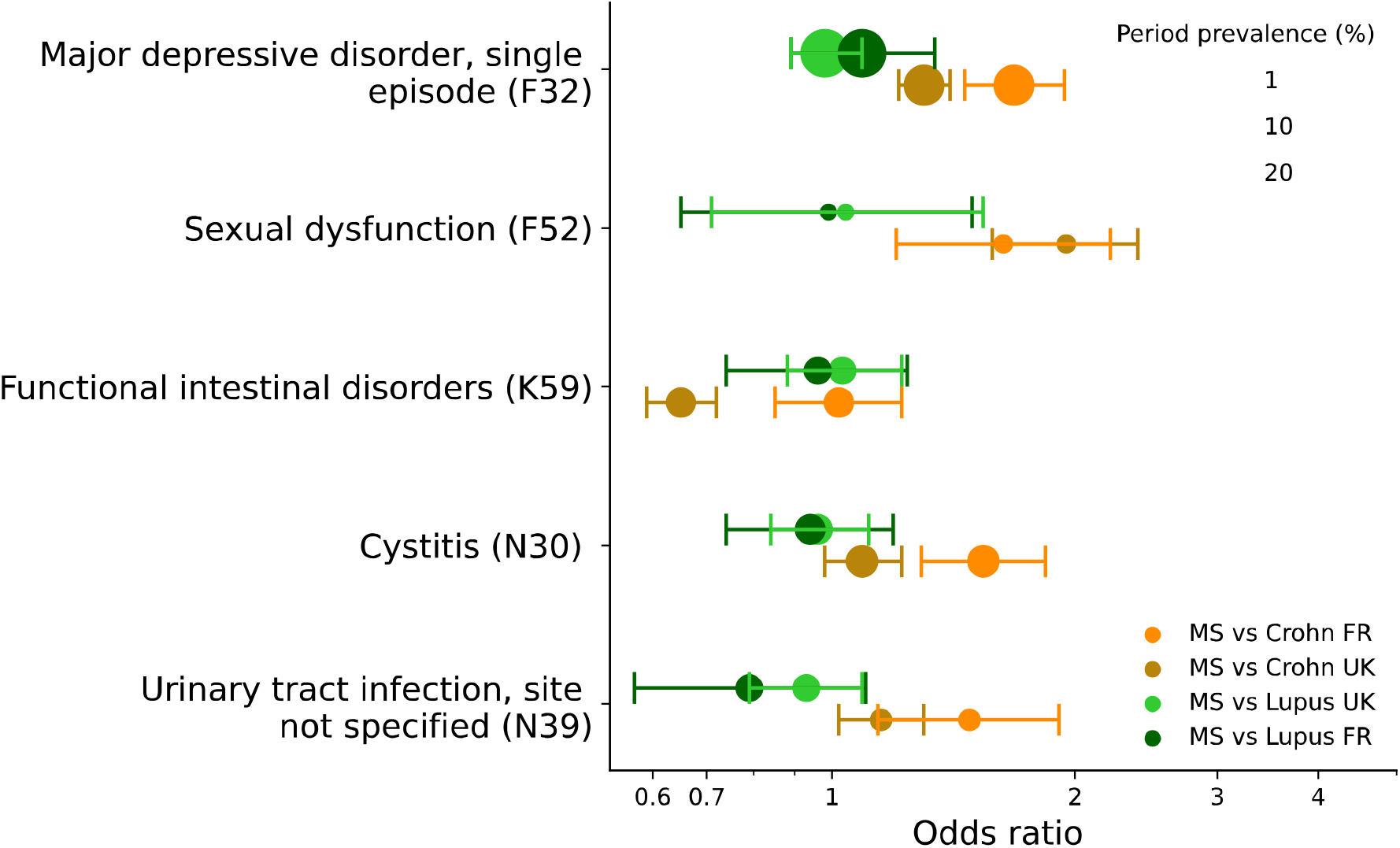
ORs for significant associations in the sensitivity analysis for five years before the index date in comparison to other auto-immune diseases. MS patients are compared to lupus and Crohn’s disease patients in both the French and UK cohorts.

### Drug prescription

We investigated changes in drug prescription over time, before and after the index date in the MS patient and control cohorts, for the health conditions identified as significantly associated with MS in the sensitivity analysis, with the exception of sexual dysfunction (Figure 3). We also investigated hormonal contraceptive use (for women only), and healthcare use, based on the mean number of visits per year (Figure 3). We found that the difference in the prevalence of antidepressant use between MS and controls was much more pronounced after than before MS diagnosis. Indeed, the period prevalence increased from 14% and 10% for MS patients and controls, respectively, in the five years before diagnosis to 37% and 19%, respectively, in the five years after MS diagnosis, with the number of MS patients using antidepressants increasing by 20% during the year of MS diagnosis. The increase in antidepressant prescription was less pronounced at the time of diagnosis for patients with lupus or Crohn’s disease.

**Figure 3.**
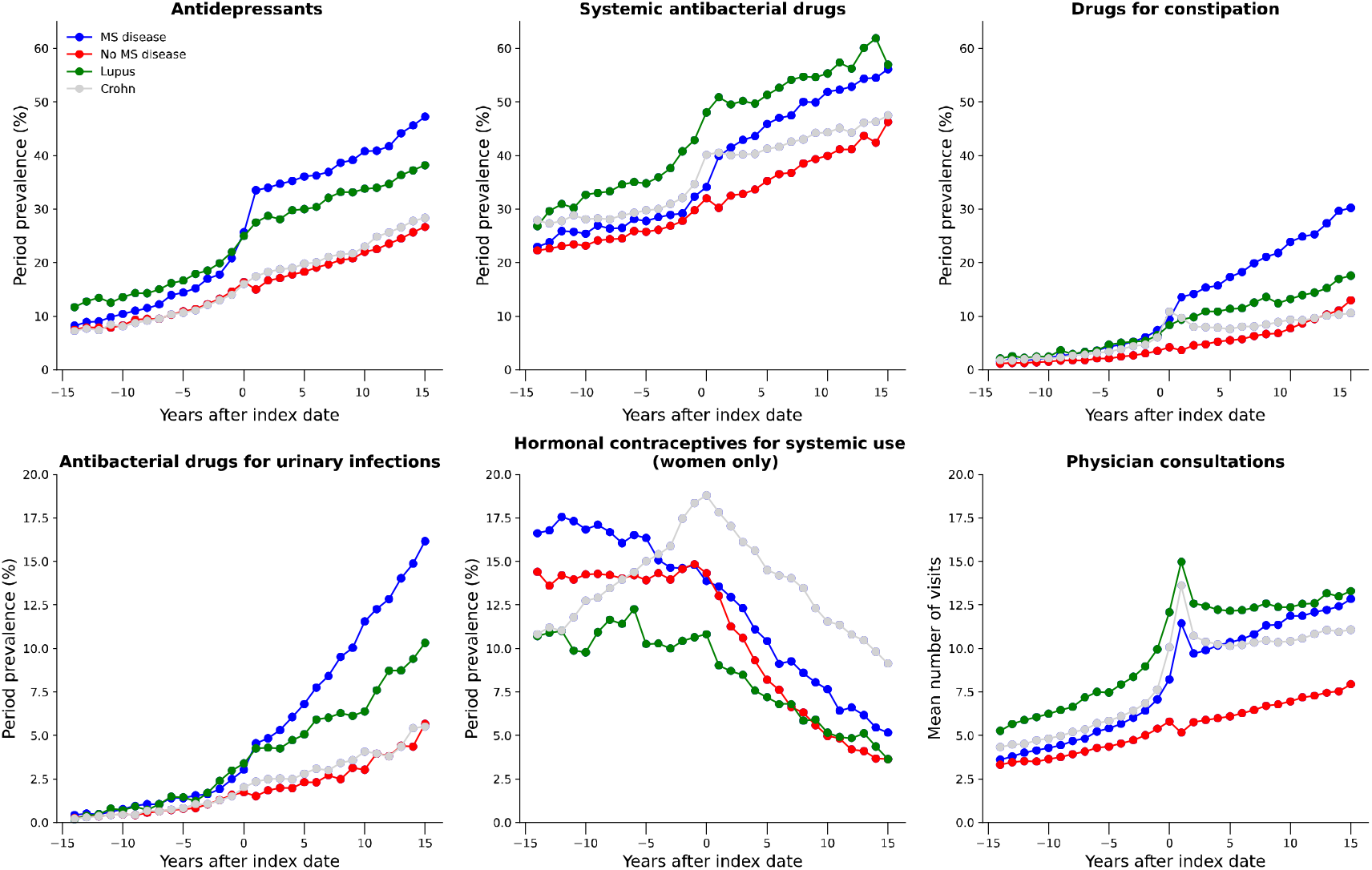
Period prevalence (%) of prescriptions associated with depression, urinary tract infections, bacterial infections, constipation, and hormonal contraceptives. The mean number of visits to the physician per year is shown on the graph at the bottom right. For a given prescription, the period prevalence is defined as the number of prescriptions observed over the period divided by the number of patients at risk during the period.

### Stratification for age, sex, and year of diagnosis

The odds ratios for autonomic symptoms (functional intestinal disorders, cystitis, and urinary tract infection) increased with increasing age at diagnosis, whereas those for depression did not vary with age. Overall, men had more pronounced pre-diagnosis presentations than women, especially for urinary tract infection (women: OR 1.26, 1.11 – 1.43; men: OR 2.39, 1.75 – 3.26). Finally, no significant differences were found for the associations with depression, constipation, cystitis, and urinary tract infections after stratification for year of diagnosis (Table 2).

## Discussion

We used two large independent primary care databases to explore pre-diagnosis health conditions associated with MS. To our knowledge, this is the first transnational data-driven study of the pre- and post-diagnosis presentations of MS. In the primary analysis, we identified a group of symptoms recorded by GPs during the five-year period leading up to MS diagnosis. However, some of these symptoms suggested that the patient had already experienced a relapse or onset of MS and, therefore, that the diagnosis of MS recorded in the database was later than the actual onset of MS.^11^ In the sensitivity analysis using the earliest symptom suggestive of MS as the index date, five conditions were significantly associated with a subsequent diagnosis of MS and were considered putative pre-diagnosis features. Our findings confirmed the reported pre-diagnosis associations for autonomic symptoms (intestinal function disorders, cystitis, and urinary tract infections) ^8,9,21,22^ and depression.^7,8,21,22^ Symptoms of sexual dysfunction have been recognized to occur at or after the onset of MS symptoms or MS diagnosis; ^23–25^ however, to our knowledge, no other study has ever reported a health condition related to sexual dysfunction as a pre-diagnosis feature of MS. The sexual dysfunctions reported were coded as a psychological disorder not caused by a disease. It remains unclear whether this health condition results from the patient’s experience of the prodrome — in which case, the psychological nature of the sexual dysfunction would be confirmed — or whether it is caused by an ongoing disease, such as medullary MS lesions, if the patient reports no motor or sensory symptoms. Other health conditions identified in prodrome studies in the primary care context, such as anxiety, fatigue, anemia, and sleep disorders, ^8,22,26^ were not significantly associated with MS diagnosis in our sensitivity study. This study is the first to investigate the possibility of an MS prodrome in two large countries, and it may have eliminated several spurious correlations.

Little is known about the similarities and differences between autoimmune diseases, particularly as concerns the nature of pre-diagnosis features. With the exception of a female preponderance, none of the observed associations was selectively associated with MS in a comparison with lupus, whereas several of these associations were found to be selectively associated with MS in a comparison with Crohn’s disease. However, these associations were weak, suggesting that depression and sexual dysfunction may not be specific for MS. Depression was the only one of these five health conditions compared between MS and Crohn’s disease in a previous study, which found no association.^11^ A previous review reported that studies of the preclinical phase of lupus are complicated by the fact that lupus cannot be diagnosed until sufficient clinical manifestations have occurred, potentially accounting for the lack of difference between the prodromal features of MS and lupus.^27^

Only one previous study has investigated whether the pre-diagnosis presentations of MS vary with age and sex ^9^. It reported higher rates of pre-diagnosis anemia in men than in women, whereas sex had no effect on the rates of pre-diagnosis fatigue and pain. Our study is the first in which the effects of age and sex on the psychiatric and autonomic pre-diagnostic features of MS were evaluated. Our findings suggest that pre-diagnostic presentations are more pronounced in men than in women. An effect of age was found only for autonomic symptoms, for which the odds ratio increased with age. Finally, the associations were weaker for diagnoses established after 2010, but this difference was not statistically significant due to the small sample size. The updated diagnostic criteria have probably decreased the time to diagnosis, as shown by the four-year decrease in the UK and the two-year decrease in France in the age at which patients are diagnosed with MS, since 2010.

For the significant associations, it remains unclear whether the conditions identified are risk factors for MS, or nonspecific early symptoms of MS. It also remains unclear whether these symptoms can be used to improve the early diagnosis of MS. We found that disclosure of the diagnosis and the progression of MS disease had a strong impact on all these associations. In large EHR databases, some time may elapse between disclosure of the diagnosis and the first record for a few patients. Given the large increase in antidepressant use after MS diagnosis, we cannot rule out the possibility that the observed association with depression can be explained by a few patients for whom the first record of MS appears several years after the first true diagnosis. Caution is therefore required in interpretation and it would probably be hasty to conclude that use of these observed prodromal associations would improve the early diagnosis of MS.

Our study has several strengths. It is the first fully agnostic study of the MS prodrome, based on data from two large databases in two different countries. This should ensure that the results are highly generalizable. This study also includes the first comparison with the period after MS diagnosis, and the first stratification for age, sex, and year of diagnosis, in line with the recommendations of previous studies.^10^ However, like most studies based on primary care databases, it is also subject to several limitations. First, no data for potential confounding factors, such as education level, ethnicity, socioeconomic status, body-mass index, or genetic information, were available. Second, the controls were also selected from primary care databases, and the control cohort did not, therefore, include individuals who never consulted a general practitioner.

## Conclusion

We performed a modern large-scale data-driven survey of a potential prodrome of MS in the primary care setting. Our findings confirm that several pre-diagnostic features of MS can be observed in large-scale electronic health record data. We also show that the prodromal presentation depends on age at diagnosis and that all pre-diagnosis health conditions associated with MS were recorded more frequently in the [0-5] years after MS diagnosis compared to the [0-5] years before MS diagnosis. Future studies are required to determine whether the detection of these health conditions can help to decrease age at diagnosis of MS, although our results suggest that they are probably not sufficiently specific to define an MS prodrome.

## Data Availability

The data used in the preparation of the article are available from the Cegedim company upon reasonable request.

