## Supplementary material for "Comparing pre- and post-diagnosis presentations of multiple sclerosis and other inflammatory diseases in primary care: an agnostic study of French and British health records"

### Supplementary materials

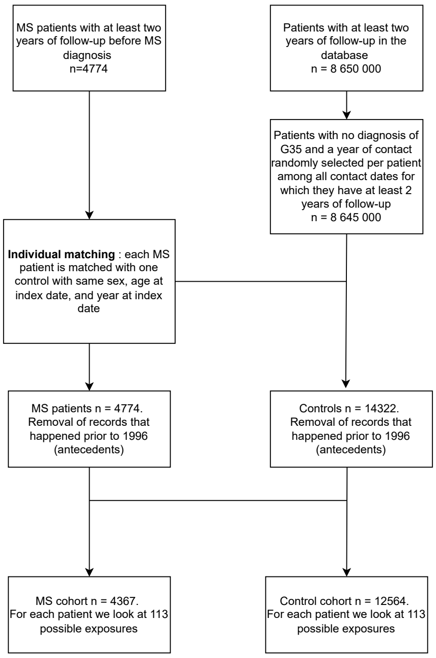


*Figure 1 Patient selection flowchart for the French study. Note : the index date for controls is taken as the last contact date recorded during tje year of contact selected randomly*

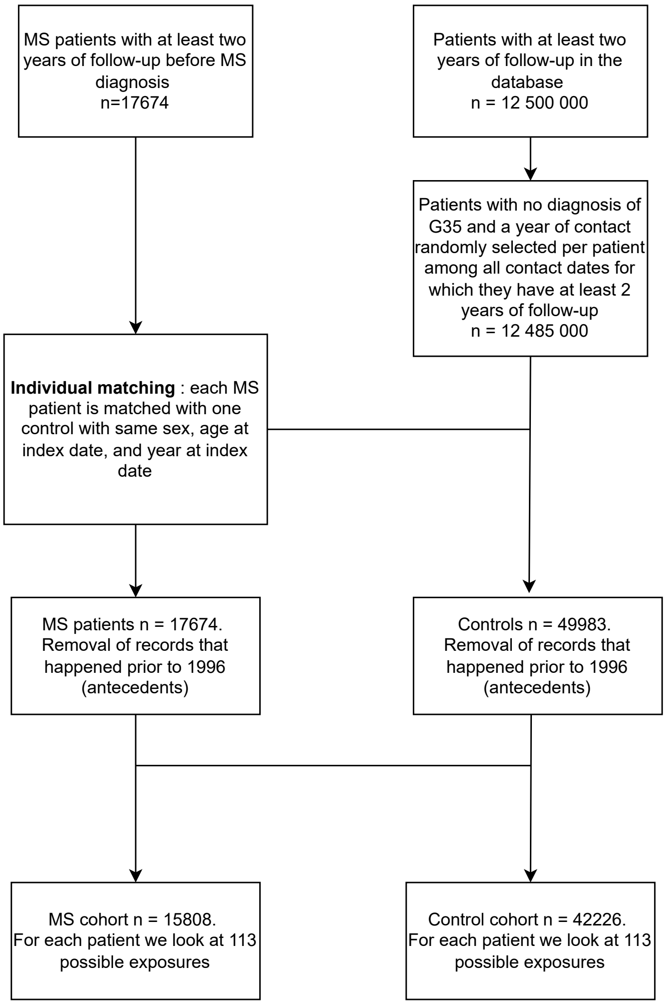


*Figure 2 Patient selection flowchart for the UK study. Note : the index date for controls is taken as the last contact date recorded during tje year of contact selected randomly*

*Table 1 Prevalence of health conditions tested in the MS populations in France and the UK at 5 years before the diagnosis. The 113 ICD10 codes that were recorded with a relative frequency of more than 0.5 % in both countries were considered.*

| | FR Controls (UK) n=42266 | UK Controls (FR) n=12564 |
|---|---|---|
| B00 : Herpesviral [herpes simplex] infections | 3.36 | 2.02 |
| B02 : Zoster | 1.73 | 0.84 |
| B07 : Viral warts | 3.11 | 0.73 |
| B34 : Viral infection of unspecified site | 2.12 | 2.08 |
| B35 : Dermatophytosis | 5.5 | 2.39 |
| B37 : Candidiasis | 5.54 | 2.63 |
| D50 : Iron deficiency anemia | 1.89 | 1.09 |
| D64 : Other anemias | 0.73 | 0.97 |
| E03 : Other hypothyroidism | 4.51 | 2.39 |
| E11 : Type 2 diabetes mellitus | 3.77 | 2.45 |
| E14 : Other diabetes | 1.71 | 0.76 |

| | | |
|---|---|---|
| E66 : Overweight and obesity | 1.48 | 1.88 |
| E78 : Disorders of lipoprotein metabolism and other lipidemias | 9.02 | 6.51 |
| F10 : Alcohol related disorders | 1.19 | 0.59 |
| F32 : Major depressive disorder, single episode | 19.65 | 10.75 |
| F40 : Phobic anxiety disorders | 0.88 | 1.19 |
| F41 : Other anxiety disorders | 8.43 | 10.27 |
| F43 : Reaction to severe stress | 1.49 | 3.3 |
| F52 : Sexual dysfunction | 1.54 | 1.96 |
| G40 : Epilepsy and recurrent seizures | 0.86 | 0.57 |
| G43 : Migraine | 4.7 | 4.9 |
| G47 : Sleep disorders | 1.92 | 6.47 |
| G56 : Carpal tunnel syndrome | 1.45 | 1.13 |
| H00 : Hordeolum and chalazion | 1.59 | 0.85 |
| H10 : Conjunctivitis | 6.02 | 4.21 |
| H60 : Otitis externa | 4.53 | 1.89 |
| H61 : Other disorders of external ear | 3.11 | 0.93 |
| H66 : Suppurative and unspecified otitis media | 2.2 | 5.1 |
| H81 : Disorders of vestibular function | 1.32 | 1.47 |
| H92 : Otalgia and effusion of ear | 2.51 | 3.01 |
| H93 : Other disorders of ear | 1.03 | 1.05 |
| I10 : Essential (primary) hypertension | 13.7 | 10.51 |
| I20 : Angina pectoris | 1.14 | 0.58 |
| I80 : Phlebitis and thrombophlebitis | 1.09 | 0.71 |
| I83 : Varicose veins of lower extremities | 1.97 | 1.14 |
| I84 : Hemorrhoids | 6.62 | 3.37 |
| J00 : Acute nasopharyngitis [common cold] | 2.71 | 24.56 |
| J01 : Acute sinusitis | 5.7 | 9.52 |
| J02 : Acute pharyngitis | 6.52 | 17.18 |
| J04 : Acute laryngitis and tracheitis | 1.8 | 11.36 |
| J11 : Influenza due to unidentified influenza virus | 2.97 | 11.58 |
| J20 : Acute bronchitis | 2.54 | 9.62 |
| J30 : Vasomotor and allergic rhinitis | 5.0 | 2.71 |
| J31 : Chronic rhinitis, nasopharyngitis and pharyngitis | 1.17 | 2.16 |
| J32 : Chronic sinusitis | 0.69 | 0.82 |
| J34 : Other and unspecified disorders of nose and nasal sinuses | 1.17 | 2.11 |
| J40 : Bronchitis | 0.6 | 3.53 |
| J45 : Asthma | 7.99 | 5.07 |
| K04 : Diseases of pulp and periapical tissues | 1.06 | 1.03 |
| K12 : Stomatitis and related lesions | 0.98 | 0.9 |
| K21 : Gastro-esophageal reflux disease | 2.76 | 4.86 |
| K29 : Gastritis and duodenitis | 1.66 | 3.42 |
| K30 : Functional dyspepsia | 3.72 | 1.1 |

| | | |
|---|---|---|
| K44 : Diaphragmatic hernia | 1.24 | 5.91 |
| K58 : Irritable bowel syndrome | 3.42 | 4.25 |
| K59 : Functional intestinal disorders | 4.37 | 5.22 |
| K62 : Other diseases of anus and rectum | 2.44 | 0.9 |
| K80 : Cholelithiasis | 0.89 | 0.85 |
| L02 : Cutaneous abscess, furuncle and carbuncle | 2.21 | 1.55 |
| L03 : Cellulitis and acute lymphangitis | 3.43 | 0.89 |
| L21 : Seborrheic dermatitis | 1.44 | 0.6 |
| L29 : Pruritus | 2.43 | 3.17 |
| L30 : Other and unspecified dermatitis | 4.74 | 4.8 |
| L40 : Psoriasis | 2.07 | 1.13 |
| L50 : Urticaria | 1.69 | 1.0 |
| L65 : Other nonscarring hair loss | 0.98 | 1.05 |
| L70 : Acne | 3.51 | 1.6 |
| L98 : Other disorders of skin and subcutaneous tissue | 4.61 | 0.94 |
| M10 : Gout | 1.11 | 0.53 |
| M13 : Other arthritis | 1.16 | 0.89 |
| M19 : Other and unspecified osteoarthritis | 3.09 | 3.77 |
| M25 : Pain in joint | 8.05 | 8.63 |
| M47 : Spondylosis | 1.3 | 1.82 |
| M51 : Thoracic, thoracolumbar, and lumbosacral intervertebral disc disorders | 0.84 | 0.93 |
| M53 : Cervicobrachial syndrome | 0.73 | 2.82 |
| M54 : Dorsalgia | 15.5 | 24.57 |
| M62 : Contracture of muscle | 0.63 | 2.09 |
| M75 : Shoulder lesions | 2.53 | 5.6 |
| M77 : Other enthesopathies | 3.31 | 6.11 |
| M79 : Pain in limb, foot / Myalgia | 14.24 | 8.77 |
| M81 : Osteoporosis without current pathological fracture | 0.96 | 0.98 |
| N30 : Cystitis | 6.99 | 6.81 |
| N39 : Urinary tract infection, site not specified | 5.36 | 2.77 |
| N64 : Other disorders of breast | 2.72 | 0.9 |
| N76 : Other inflammation of vagina and vulva | 1.25 | 1.17 |
| N91 : Absent, scanty and rare menstruation | 2.23 | 1.44 |
| N92 : Excessive, frequent and irregular menstruation | 7.58 | 1.72 |
| N94 : Pain and other conditions associated with female genital organs and menstrual cycle | 2.33 | 1.52 |
| N95 : Menopausal and other perimenopausal disorders | 4.91 | 3.13 |
| R05 : Cough | 0.83 | 16.59 |
| R06 : Abnormalities of breathing | 1.36 | 1.93 |
| R07 : Pain in throat and chest | 2.0 | 3.92 |
| R10 : Abdominal and pelvic pain | 4.93 | 12.45 |
| R20 : Paresthesia of skin | 0.86 | 0.82 |
| R21 : Rash and other nonspecific skin eruption | 2.35 | 1.13 |

| | | |
|---|---|---|
| R25 : Abnormal involuntary movements | 1.25 | 1.33 |
| R42 : Dizziness and giddiness | 3.95 | 3.31 |
| R51 : Headache | 2.0 | 7.23 |
| R52 : Pain, unspecified | 0.99 | 4.52 |
| R53 : Malaise and fatigue | 1.64 | 12.53 |
| R55 : Syncope and collapse | 0.94 | 0.85 |
| R59 : Enlarged lymph nodes | 0.65 | 1.54 |
| R60 : Edema, not elsewhere classified | 0.92 | 1.32 |
| R63 : Abnormal weight loss or gain | 0.68 | 1.32 |
| S83 : Dislocation and sprain of joints and ligaments of knee | 0.78 | 0.87 |
| S93 : Dislocation and sprain of joints and ligaments at ankle; foot and toe level | 1.32 | 2.39 |
| T14 : Injury of unspecified body region | 3.55 | 8.27 |
| T78 : Adverse effects, not elsewhere classified | 1.56 | 8.34 |
| Z01 : Encounter for other special examination without complaint, suspected or reported diagnosis | 0.6 | 1.63 |
| Z02 : Encounter for administrative examination | 3.61 | 11.89 |
| Z13 : Encounter for screening for other diseases and disorders | 0.75 | 2.09 |
| Z30 : Encounter for contraceptive management | 14.1 | 8.36 |
| Z76 : Persons encountering health services in other circumstances | 1.5 | 10.29 |

*Table 2 Prescriptions linked to diagnoses in the French database. Prescriptions are deemed as linked when they were given to more than 80 different people, and given in more than one out of three times for a particular diagnosis/symptom*

| | |
|---|---|
| Ispaghula (psylla seeds) (A06AC01) | Other functional intestinal disorders (K59) |
| Macrogol (A06AD15) | Other functional intestinal disorders (K59) |
| Macrogol, combinations (A06AD65) | Other functional intestinal disorders (K59) |
| Enema combinations (A06AG20) | Other functional intestinal disorders (K59) |
| Carbon dioxide producing drugs (A06AX02) | Other functional intestinal disorders (K59) |
| Amphotericin B (A07AA07) | Candidiasis (B37) |
| Insulin glargine (A10AE04) | Type 2 diabetes mellitus (E11) |
| Metformin (A10BA02) | Type 2 diabetes mellitus (E11) |
| Gliclazide (A10BB09) | Type 2 diabetes mellitus (E11) |
| Glimepiride (A10BB12) | Type 2 diabetes mellitus (E11) |
| Metformin and sitagliptin (A10BD07) | Type 2 diabetes mellitus (E11) |
| Sitagliptin (A10BH01) | Type 2 diabetes mellitus (E11) |
| Repaglinide (A10BX02) | Type 2 diabetes mellitus (E11) |
| Vitamins, other combinations (A11JC) | Other nonscarring hair loss (L65) |
| Tranexamic acid (B02AA02) | Excessive, frequent and irregular menstruation (N92) |
| Hydrochlorothiazide (C03AA03) | Essential (primary) hypertension (I10) |
| Indapamide (C03BA11) | Essential (primary) hypertension (I10) |

| | |
|---|---|
| Spironolactone (C03DA01) | Essential (primary) hypertension (I10) |
| Other agents for treatment of hemorrhoids and anal fissures for topical use in ATC (C05AX) | Hemorrhoids (I84) |
| Other preparations, combinations (C05AX03) | Hemorrhoids (I84) |
| Rutoside, combinations (C05CA51) | Other disorders of veins (I87) |
| Atenolol (C07AB03) | Essential (primary) hypertension (I10) |
| Acebutolol (C07AB04) | Essential (primary) hypertension (I10) |
| Bisoprolol (C07AB07) | Essential (primary) hypertension (I10) |
| Celiprolol (C07AB08) | Essential (primary) hypertension (I10) |
| Nebivolol (C07AB12) | Essential (primary) hypertension (I10) |
| Amlodipine (C08CA01) | Essential (primary) hypertension (I10) |
| Nicardipine (C08CA04) | Essential (primary) hypertension (I10) |
| Lercanidipine (C08CA13) | Essential (primary) hypertension (I10) |
| Perindopril (C09AA04) | Essential (primary) hypertension (I10) |
| Ramipril (C09AA05) | Essential (primary) hypertension (I10) |
| Perindopril and diuretics (C09BA04) | Essential (primary) hypertension (I10) |
| Losartan (C09CA01) | Essential (primary) hypertension (I10) |
| Valsartan (C09CA03) | Essential (primary) hypertension (I10) |
| Irbesartan (C09CA04) | Essential (primary) hypertension (I10) |
| Candesartan (C09CA06) | Essential (primary) hypertension (I10) |
| Telmisartan (C09CA07) | Essential (primary) hypertension (I10) |
| Olmesartan medoxomil (C09CA08) | Essential (primary) hypertension (I10) |
| Losartan and diuretics (C09DA01) | Essential (primary) hypertension (I10) |
| Valsartan and diuretics (C09DA03) | Essential (primary) hypertension (I10) |
| Irbesartan and diuretics (C09DA04) | Essential (primary) hypertension (I10) |
| Valsartan and amlodipine (C09DB01) | Essential (primary) hypertension (I10) |
| Simvastatin (C10AA01) | Disorders of lipoprotein metabolism and other lipidemias (E78) |
| Pravastatin (C10AA03) | Disorders of lipoprotein metabolism and other lipidemias (E78) |
| Fluvastatin (C10AA04) | Disorders of lipoprotein metabolism and other lipidemias (E78) |
| Atorvastatin (C10AA05) | Disorders of lipoprotein metabolism and other lipidemias (E78) |
| Rosuvastatin (C10AA07) | Disorders of lipoprotein metabolism and other lipidemias (E78) |
| Fenofibrate (C10AB05) | Disorders of lipoprotein metabolism and other lipidemias (E78) |
| Ezetimibe (C10AX09) | Disorders of lipoprotein metabolism and other lipidemias (E78) |
| Amorolfine (D01AE16) | Dermatophytosis (B35) |
| Terbinafine (D01BA02) | Dermatophytosis (B35) |
| Calcipotriol, combinations (D05AX52) | Psoriasis (L40) |

| | |
|---|---|
| Aciclovir (D06BB03) | Herpesviral [herpes simplex] infections (B00) |
| Metronidazole (D06BX01) | Rosacea (L71) |
| Benzoyl peroxide (D10AE01) | Acne (L70) |
| Erythromycin (D10AF02) | Acne (L70) |
| Wart and anti-corn preparations (D11AF) | Viral warts (B07) |
| Other gynecologicals in ATC (G02CX) | Menopausal and other perimenopausal disorders (N95) |
| Levonorgestrel and ethinylestradiol (G03AA07) | Encounter for contraceptive management (Z30) |
| Desogestrel and ethinylestradiol (G03AA09) | Encounter for contraceptive management (Z30) |
| Gestodene and ethinylestradiol (G03AA10) | Encounter for contraceptive management (Z30) |
| Drospirenone and ethinylestradiol (G03AA12) | Encounter for contraceptive management (Z30) |
| Levonorgestrel and ethinylestradiol (G03AB03) | Encounter for contraceptive management (Z30) |
| Levonorgestrel (G03AC03) | Encounter for contraceptive management (Z30) |
| Desogestrel (G03AC09) | Encounter for contraceptive management (Z30) |
| Estradiol (G03CA03) | Menopausal and other perimenopausal disorders (N95) |
| Cyproterone and estrogen (G03HB01) | Encounter for contraceptive management (Z30) |
| Tadalafil (G04BE08) | Sexual dysfunction not caused by organic disorder or disease (F52) |
| Tamsulosin (G04CA02) | Benign prostatic hyperplasia (N40) |
| Levothyroxine sodium (H03AA01) | Other hypothyroidism (E03) |
| Pivmecillinam (J01CA08) | Cystitis (N30) |
| Phenoxymethylpenicillin (J01CE02) | Acute pharyngitis (J02) |
| Norfloxacin (J01MA06) | Cystitis (N30) |
| Moxifloxacin (J01MA14) | Acute sinusitis (J01) |
| Nitrofurantoin (J01XE01) | Cystitis (N30) |
| Fosfomycin (J01XX01) | Cystitis (N30) |
| Aciclovir (J05AB01) | Herpesviral [herpes simplex] infections (B00) |
| Oseltamivir (J05AH02) | Influenza due to unidentified influenza virus (J11) |
| Pneumococcus, purified polysaccharides antigen (J07AL01) | Need for immunization against other single infectious diseases (Z26) |
| Tetanus toxoid (J07AM01) | Need for immunization against other single infectious diseases (Z26) |
| Typhoid, purified polysaccharide antigen (J07AP03) | Need for immunization against other single infectious diseases (Z26) |
| Hepatitis B, purified antigen (J07BC01) | Need for immunization against other single infectious diseases (Z26) |
| Hepatitis A, inactivated, whole virus (J07BC02) | Need for immunization against other single infectious diseases (Z26) |

| | |
|---|---|
| Measles, combinations with mumps and rubella, live attenuated (J07BD52) | Need for immunization against other single infectious diseases (Z26) |
| Papillomavirus (human types 6, 11, 16, 18) (J07BM01) | Need for immunization against other single infectious diseases (Z26) |
| Diphtheria-poliomyelitis-tetanus (J07CA01) | Need for immunization against other single infectious diseases (Z26) |
| Diphtheria-pertussis-poliomyelitis-tetanus (J07CA02) | Need for immunization against other single infectious diseases (Z26) |
| Meloxicam (M01AC06) | Dorsalgia (M54) |
| Methocarbamol (M03BA03) | Dorsalgia (M54) |
| Colchicine (M04AC01) | Gout (M10) |
| Risedronic acid (M05BA07) | Osteoporosis without current pathological fracture (M81) |
| Sumatriptan (N02CC01) | Migraine (G43) |
| Naratriptan (N02CC02) | Migraine (G43) |
| Zolmitriptan (N02CC03) | Migraine (G43) |
| Almotriptan (N02CC05) | Migraine (G43) |
| Eletriptan (N02CC06) | Migraine (G43) |
| Frovatriptan (N02CC07) | Migraine (G43) |
| Loprazolam (N05CD11) | Sleep disorders (G47) |
| Fluoxetine (N06AB03) | Major depressive disorder, single episode (F32) |
| Citalopram (N06AB04) | Major depressive disorder, single episode (F32) |
| Paroxetine (N06AB05) | Major depressive disorder, single episode (F32) |
| Sertraline (N06AB06) | Major depressive disorder, single episode (F32) |
| Escitalopram (N06AB10) | Major depressive disorder, single episode (F32) |
| Mianserin (N06AX03) | Major depressive disorder, single episode (F32) |
| Mirtazapine (N06AX11) | Major depressive disorder, single episode (F32) |
| Venlafaxine (N06AX16) | Major depressive disorder, single episode (F32) |
| Duloxetine (N06AX21) | Major depressive disorder, single episode (F32) |
| Varenicline (N07BA03) | Nicotine dependence (F17) |
| Betahistine (N07CA01) | Dizziness and giddiness (R42) |
| Proguanil, combinations (P01BB51) | Persons encountering health services for other counseling and medical advice, not elsewhere classified (Z71) |
| Ivermectin (P02CF01) | Scabies (B86) |
| Benzyl benzoate (P03AX01) | Scabies (B86) |
| Dexamethasone, combinations (R01AD53) | Acute nasopharyngitis [common cold] (J00) |
| Other nasal preparation combinations in ATC (R01AX30) | Acute nasopharyngitis [common cold] (J00) |
| Pseudoephedrine (R01BA02) | Acute nasopharyngitis [common cold] (J00) |
| Fusafungine (R02AB03) | Acute nasopharyngitis [common cold] (J00) |
| Terbutaline (R03AC03) | Asthma (J45) |
| Formoterol (R03AC13) | Asthma (J45) |
| Salmeterol and fluticasone (R03AK06) | Asthma (J45) |

| | |
|---|---|
| Montelukast (R03DC03) | Asthma (J45) |
| OTHER COLD PREPARATIONS in ATC (R05X) | Acute nasopharyngitis [common cold] (J00) |
| Tobramycin (S01AA12) | Conjunctivitis (H10) |
| Fusidic acid (S01AA13) | Conjunctivitis (H10) |
| Azithromycin (S01AA26) | Conjunctivitis (H10) |
| Sodium borate (S01AX07) | Conjunctivitis (H10) |
| Indifferent preparations, otologicals (S02DC) | Other disorders of external ear (H61) |

*Table 3 Health conditions suggestive of a demyelinating claim*

| |
|---|
| Encephalitis, myelitis, encephalomyelitis (G04, G05) |
| Sequelae of inflammatory diseases of central nervous system (G09) |
| Demyelinating diseases of the CNS (G36, G37) |
| Retrobulbar neuritis and other disorders of the optic nerve and the visual pathways (H46, H47, H48) |

*Table 4 Health conditions suggestive of an MS relapse and positively associated with a future diagnosis of MS in either the French or British database.*

| |
|---|
| Paresthesia of skin (R20) |
| Tetany (R29) |
| Dizziness and giddiness (R42) |
| Abnormal and involuntary movement (R25) |
| Carpal tunnel syndrome (G56) |
| Disorders of vestibular function (H81) |
| Visual disturbances (H53) |
| Contracture of muscle (M62) |

*Table 5 ORs for all health troubles in over 0.5% of patients and controls individually associated with a future symptom specific of MS at 5 years before the index date. In the primary analysis we used the first recorded demyelinating event as the index date, while in the sensitivity analysis the first symptom suggestive of MS is used as the index date. .* cannot be*

| | Primary analysis | | | | Sensitivity analysis | | | |
|---|---|---|---|---|---|---|---|---|
| | FR | | UK | | FR | | UK | |
| | OR (Adjusted 95% CI) | Adjusted p value | OR (Adjusted 95% CI) | Adjusted p value | OR (Adjusted 95% CI) | Adjusted p value | OR (Adjusted 95% CI) | Adjusted p value |
| B00 : Herpesviral [herpes simplex] infections | 1.36 (0.89 - 2.08) | 1.138 | 1.02 (0.84 - 1.24) | 136.872 | 1.06 (0.66 - 1.73) | 99.044 | 1.0 (0.80 - 1.24) | 155.186 |
| B02 : Zoster | 2.59 (1.49 - 4.51) | <0.0001 | 0.93 (0.70 - 1.23) | 56.949 | 1.32 (0.66 - 2.64) | 20.692 | 0.85 (0.62 - 1.16) | 7.945 |

| | | | | | | | | |
|---|---|---|---|---|---|---|---|---|
| B07 : Viral warts | 1.51 (0.76 - 2.98) | 3.902 | 0.74 (0.59 - 0.93) | 0.0001 | 1.13 (0.52 - 2.46) | 88.512 | 0.82 (0.65 - 1.03) | 0.159 |
| B34 : Viral infection of unspecified site | 1.4 (0.92 - 2.13) | 0.38 | 1.0 (0.77 - 1.28) | 179.35 | 1.4 (0.89 - 2.20) | 0.731 | 0.96 (0.73 - 1.25) | 88.167 |
| B35 : Dermatophytosis | 1.5 (1.03 - 2.20) | 0.009 | 0.89 (0.76 - 1.05) | 1.422 | 1.29 (0.84 - 1.98) | 4.084 | 0.88 (0.74 - 1.05) | 1.315 |
| B37 : Candidiasis | 1.58 (1.10 - 2.27) | 0.0003 | 0.88 (0.74 - 1.03) | 0.397 | 1.35 (0.89 - 2.04) | 0.958 | 0.82 (0.69 - 0.98) | 0.006 |
| D50 : Iron deficiency anemia | 1.82 (1.07 - 3.10) | 0.004 | 0.8 (0.60 - 1.06) | 0.537 | 1.71 (0.97 - 3.03) | 0.059 | 0.73 (0.53 - 1.00) | 0.03 |
| D64 : Other anemias | 1.3 (0.70 - 2.42) | 19.406 | 1.04 (0.69 - 1.59) | 131.168 | 1.02 (0.49 - 2.11) | 144.633 | 1.0 (0.63 - 1.58) | 161.172 |
| E03 : Other hypothyroidism | 1.35 (0.90 - 2.00) | 0.771 | 1.0 (0.84 - 1.19) | 180.453 | 1.28 (0.82 - 1.99) | 5.702 | 1.01 (0.84 - 1.21) | 147.701 |
| E11 : Type 2 diabetes mellitus | 1.56 (1.07 - 2.28) | 0.001 | 0.76 (0.62 - 0.93) | <0.0001 | 1.27 (0.82 - 1.96) | 6.208 | 0.78 (0.62 - 0.97) | 0.003 |
| E14 : Other diabetes | 0.9 (0.41 - 1.98) | 107.57 | 0.85 (0.63 - 1.14) | 5.996 | 0.76 (0.31 - 1.87) | 38.822 | 0.89 (0.65 - 1.22) | 24.111 |
| E66 : Overweight and obesity | 1.25 (0.79 - 1.99) | 10.828 | 0.73 (0.53 - 1.03) | 0.076 | 1.26 (0.78 - 2.04) | 10.656 | 0.66 (0.46 - 0.96) | 0.005 |
| E78 : Disorders of lipoprotein metabolism and other lipidemias | 1.27 (0.99 - 1.64) | 0.053 | 0.85 (0.75 - 0.97) | 0.0003 | 1.07 (0.81 - 1.42) | 55.88 | 0.8 (0.69 - 0.92) | <0.0001 |
| F10 : Alcohol related disorders | 1.38 (0.63 - 3.05) | 20.963 | 0.55 (0.36 - 0.84) | <0.0001 | * | * | 0.51 (0.32 - 0.81) | <0.0001 |
| F32 : Major depressive disorder, single episode | 1.84 (1.52 - 2.21) | <0.0001 | 1.24 (1.14 - 1.36) | <0.0001 | 1.62 (1.32 - 1.98) | <0.0001 | 1.22 (1.11 - 1.34) | <0.0001 |
| F40 : Phobic anxiety disorders | 1.84 (1.09 - 3.09) | 0.001 | 0.87 (0.58 - 1.31) | 38.248 | 1.56 (0.92 - 2.66) | 0.233 | 0.73 (0.45 - 1.16) | 1.632 |
| F41 : Other anxiety disorders | 1.57 (1.29 - 1.91) | <0.0001 | 0.85 (0.74 - 0.98) | 0.001 | 1.4 (1.13 - 1.73) | <0.0001 | 0.78 (0.67 - 0.91) | <0.0001 |

| | | | | | | | | |
|---|---|---|---|---|---|---|---|---|
| F43 : Reaction to severe stress | 1.69 (1.23 - 2.32) | <0.0001 | 0.83 (0.60 - 1.15) | 5.179 | 1.47 (1.05 - 2.07) | 0.003 | 0.8 (0.58 - 1.12) | 2.049 |
| F52 : Sexual dysfunction | 1.92 (1.29 - 2.86) | <0.0001 | 1.54 (1.19 - 1.99) | <0.0001 | 1.69 (1.10 - 2.62) | 0.0008 | 1.47 (1.11 - 1.95) | <0.0001 |
| G40 : Epilepsy and recurrent seizures | 2.54 (1.30 - 4.97) | <0.0001 | 1.43 (1.00 - 2.04) | 0.021 | * | * | 1.46 (1.00 - 2.12) | 0.022 |
| G43 : Migraine | 1.55 (1.18 - 2.03) | <0.0001 | 1.12 (0.95 - 1.32) | 1.549 | 1.21 (0.89 - 1.65) | 3.224 | 1.07 (0.89 - 1.28) | 26.972 |
| G47 : Sleep disorders | 1.64 (1.30 - 2.08) | <0.0001 | 1.07 (0.82 - 1.39) | 62.234 | 1.36 (1.04 - 1.78) | 0.002 | 1.01 (0.76 - 1.34) | 150.937 |
| G56 : Carpal tunnel syndrome | 2.64 (1.64 - 4.22) | <0.0001 | 1.26 (0.95 - 1.67) | 0.304 | * | * | * | * |
| H00 : Hordeolum and chalazion | 2.53 (1.47 - 4.36) | <0.0001 | 0.92 (0.68 - 1.23) | 49.061 | 2.08 (1.12 - 3.83) | 0.001 | 0.91 (0.66 - 1.26) | 45.628 |
| H10 : Conjunctivitis | 1.42 (1.05 - 1.91) | 0.002 | 0.85 (0.73 - 1.00) | 0.026 | 1.28 (0.93 - 1.77) | 0.514 | 0.82 (0.70 - 0.98) | 0.003 |
| H60 : Otitis externa | 1.45 (0.94 - 2.24) | 0.184 | 0.85 (0.71 - 1.02) | 0.146 | 1.49 (0.93 - 2.37) | 0.195 | 0.76 (0.62 - 0.93) | <0.0001 |
| H61 : Other disorders of external ear | 1.58 (0.87 - 2.85) | 0.576 | 1.0 (0.81 - 1.23) | 179.814 | 1.28 (0.66 - 2.50) | 25.77 | 0.94 (0.75 - 1.17) | 40.928 |
| H66 : Suppurative and unspecified otitis media | 1.45 (1.11 - 1.91) | <0.0001 | 0.87 (0.68 - 1.13) | 9.118 | 1.32 (0.98 - 1.76) | 0.054 | 0.93 (0.71 - 1.22) | 51.595 |
| H81 : Disorders of vestibular function | 2.22 (1.44 - 3.43) | <0.0001 | 1.49 (1.13 - 1.97) | <0.0001 | * | * | * | * |
| H92 : Otalgia and effusion of ear | 1.59 (1.13 - 2.23) | <0.0001 | 0.9 (0.71 - 1.14) | 15.192 | 1.5 (1.03 - 2.18) | 0.007 | 0.8 (0.62 - 1.04) | 0.24 |
| H93 : Other disorders of ear, not elsewhere classified | 1.38 (0.77 - 2.46) | 6.133 | 1.02 (0.71 - 1.45) | 163.09 | 0.87 (0.42 - 1.82) | 76.415 | 0.78 (0.51 - 1.20) | 4.484 |
| I10 : Essential (primary) hypertension | 1.28 (1.05 - 1.57) | 0.0006 | 0.79 (0.70 - 0.88) | <0.0001 | 1.09 (0.87 - 1.37) | 23.439 | 0.78 (0.69 - 0.88) | <0.0001 |

| | | | | | | | | |
|---|---|---|---|---|---|---|---|---|
| I20 : Angina pectoris | 1.15 (0.49 - 2.70) | 93.365 | 0.86 (0.60 - 1.24) | 23.269 | 0.98 (0.38 - 2.52) | 149.723 | 0.78 (0.53 - 1.16) | 3.039 |
| I80 : Phlebitis and thrombophlebitis | 1.65 (0.84 - 3.24) | 0.817 | 0.93 (0.65 - 1.33) | 83.357 | 1.31 (0.57 - 3.01) | 34.746 | 0.92 (0.62 - 1.35) | 63.434 |
| I83 : Varicose veins of lower extremities | 1.47 (0.84 - 2.55) | 1.439 | 0.67 (0.50 - 0.91) | <0.0001 | 1.32 (0.72 - 2.41) | 13.17 | 0.68 (0.49 - 0.93) | 0.0006 |
| I84 : Hemorrhoids | 1.46 (1.05 - 2.04) | 0.002 | 0.82 (0.70 - 0.96) | 0.0002 | 1.28 (0.89 - 1.83) | 1.68 | 0.81 (0.68 - 0.95) | 0.0001 |
| J00 : Acute nasopharyngitis [common cold] | 1.4 (1.20 - 1.62) | <0.0001 | 1.02 (0.82 - 1.27) | 132.286 | 1.3 (1.12 - 1.52) | <0.0001 | 0.97 (0.77 - 1.23) | 105.348 |
| J01 : Acute sinusitis | 1.48 (1.20 - 1.82) | <0.0001 | 0.91 (0.78 - 1.07) | 6.206 | 1.39 (1.11 - 1.72) | <0.0001 | 0.91 (0.77 - 1.08) | 6.188 |
| J02 : Acute pharyngitis | 1.35 (1.14 - 1.59) | <0.0001 | 0.83 (0.71 - 0.96) | 0.0004 | 1.32 (1.11 - 1.57) | <0.0001 | 0.81 (0.69 - 0.96) | 0.0004 |
| J04 : Acute laryngitis and tracheitis | 1.36 (1.12 - 1.65) | <0.0001 | 0.94 (0.71 - 1.24) | 68.337 | 1.3 (1.05 - 1.59) | 0.0003 | 0.93 (0.69 - 1.25) | 54.216 |
| J11 : Influenza due to unidentified influenza virus | 1.31 (1.08 - 1.60) | <0.0001 | 0.87 (0.70 - 1.09) | 3.395 | 1.25 (1.02 - 1.54) | 0.005 | 0.86 (0.68 - 1.09) | 2.813 |
| J20 : Acute bronchitis | 1.46 (1.19 - 1.80) | <0.0001 | 0.84 (0.65 - 1.07) | 1.088 | 1.41 (1.14 - 1.75) | <0.0001 | 0.74 (0.56 - 0.97) | 0.003 |
| J30 : Vasomotor and allergic rhinitis | 1.42 (0.98 - 2.06) | 0.048 | 0.7 (0.58 - 0.85) | <0.0001 | 1.39 (0.93 - 2.08) | 0.288 | 0.72 (0.59 - 0.88) | <0.0001 |
| J31 : Chronic rhinitis, nasopharyngitis and pharyngitis | 1.44 (0.96 - 2.18) | 0.123 | 0.94 (0.67 - 1.33) | 96.375 | 1.3 (0.85 - 2.00) | 3.034 | 0.75 (0.51 - 1.11) | 0.97 |
| J32 : Chronic sinusitis | 1.45 (0.75 - 2.82) | 5.796 | 0.77 (0.47 - 1.24) | 6.401 | 1.25 (0.62 - 2.53) | 37.437 | * | * |
| J34 : Other and unspecified disorders of nose and nasal sinuses | 1.11 (0.71 - 1.73) | 66.616 | 0.81 (0.57 - 1.17) | 5.449 | 1.08 (0.66 - 1.75) | 87.726 | 0.81 (0.55 - 1.18) | 5.615 |

| | | | | | | | | |
|---|---|---|---|---|---|---|---|---|
| J40 : Bronchitis, not specified as acute or chronic | 1.51 (1.10 - 2.07) | 0.0001 | 1.18 (0.75 - 1.85) | 32.078 | 1.37 (0.96 - 1.96) | 0.126 | 1.18 (0.75 - 1.85) | 27.603 |
| J45 : Asthma | 1.38 (1.05 - 1.83) | 0.001 | 0.81 (0.70 - 0.93) | <0.0001 | 1.28 (0.95 - 1.73) | 0.266 | 0.83 (0.72 - 0.97) | 0.0007 |
| K04 : Diseases of pulp and periapical tissues | 1.6 (0.91 - 2.80) | 0.275 | 0.88 (0.61 - 1.28) | 36.275 | 1.47 (0.79 - 2.76) | 3.097 | 0.8 (0.54 - 1.20) | 6.054 |
| K12 : Stomatitis and related lesions | 1.11 (0.56 - 2.18) | 99.787 | 0.77 (0.51 - 1.14) | 2.009 | 1.03 (0.50 - 2.12) | 138.059 | 0.76 (0.50 - 1.17) | 2.639 |
| K21 : Gastro-esophageal reflux disease | 1.26 (0.94 - 1.68) | 0.48 | 0.74 (0.58 - 0.94) | 0.0005 | 1.1 (0.80 - 1.52) | 40.895 | 0.7 (0.53 - 0.92) | <0.0001 |
| K29 : Gastritis and duodenitis | 1.54 (1.11 - 2.12) | <0.0001 | 0.75 (0.55 - 1.02) | 0.081 | 1.26 (0.88 - 1.79) | 2.498 | 0.67 (0.47 - 0.95) | 0.003 |
| K30 : Functional dyspepsia | 1.23 (0.68 - 2.23) | 31.971 | 0.77 (0.63 - 0.95) | 0.0005 | 0.9 (0.44 - 1.82) | 89.074 | 0.72 (0.58 - 0.91) | <0.0001 |
| K44 : Diaphragmatic hernia | 1.42 (1.10 - 1.83) | <0.0001 | 0.67 (0.46 - 0.98) | 0.011 | 1.31 (1.00 - 1.72) | 0.027 | 0.67 (0.45 - 1.01) | 0.042 |
| K58 : Irritable bowel syndrome | 1.43 (1.06 - 1.93) | 0.0007 | 0.82 (0.66 - 1.01) | 0.068 | 1.42 (1.04 - 1.94) | 0.003 | 0.82 (0.65 - 1.03) | 0.189 |
| K59 : Functional intestinal disorders | 1.9 (1.47 - 2.44) | <0.0001 | 1.58 (1.35 - 1.84) | <0.0001 | 1.66 (1.27 - 2.17) | <0.0001 | 1.5 (1.27 - 1.78) | <0.0001 |
| K62 : Other diseases of anus and rectum | 1.4 (0.75 - 2.61) | 7.17 | 0.86 (0.68 - 1.10) | 4.413 | 1.11 (0.53 - 2.33) | 95.581 | 0.77 (0.58 - 1.01) | 0.05 |
| K80 : Cholelithiasis | 1.35 (0.69 - 2.63) | 14.93 | 0.81 (0.54 - 1.22) | 9.522 | 1.33 (0.67 - 2.66) | 17.945 | 0.74 (0.47 - 1.19) | 2.76 |
| L02 : Cutaneous abscess, furuncle and carbuncle | 1.5 (0.93 - 2.40) | 0.201 | 0.84 (0.65 - 1.09) | 2.209 | 1.37 (0.80 - 2.34) | 4.201 | 0.82 (0.62 - 1.09) | 1.334 |
| L03 : Cellulitis and acute lymphangitis | 1.78 (0.99 - 3.20) | 0.036 | 1.01 (0.83 - 1.23) | 168.153 | 1.57 (0.82 - 3.03) | 1.377 | 0.92 (0.74 - 1.15) | 26.529 |
| L21 : Seborrheic dermatitis | 1.67 (0.81 - 3.45) | 1.203 | 0.86 (0.63 - 1.19) | 15.33 | * | * | 0.88 (0.63 - 1.24) | 27.025 |

| | | | | | | | | |
|---|---|---|---|---|---|---|---|---|
| L29 : Pruritus | 1.3 (0.91 - 1.85) | 0.865 | 0.99 (0.78 - 1.25) | 166.652 | 1.16 (0.79 - 1.71) | 22.708 | 0.85 (0.65 - 1.11) | 3.34 |
| L30 : Other and unspecified dermatitis | 1.43 (1.08 - 1.89) | 0.0002 | 1.0 (0.84 - 1.18) | 172.742 | 1.37 (1.01 - 1.87) | 0.016 | 1.0 (0.84 - 1.20) | 150.669 |
| L40 : Psoriasis | 1.97 (1.18 - 3.27) | <0.0001 | 0.92 (0.71 - 1.20) | 45.939 | 1.44 (0.80 - 2.61) | 2.915 | 0.91 (0.69 - 1.20) | 31.244 |
| L50 : Urticaria | 1.83 (1.05 - 3.18) | 0.006 | 0.83 (0.61 - 1.12) | 3.23 | 1.84 (1.02 - 3.30) | 0.014 | 0.71 (0.51 - 1.00) | 0.031 |
| L65 : Other nonscarring hair loss | 1.84 (1.07 - 3.16) | 0.003 | 0.68 (0.45 - 1.04) | 0.089 | 1.92 (1.06 - 3.49) | 0.006 | 0.72 (0.46 - 1.12) | 0.871 |
| L70 : Acne | 1.86 (1.20 - 2.89) | <0.0001 | 0.79 (0.64 - 0.98) | 0.006 | 1.72 (1.08 - 2.75) | 0.002 | 0.77 (0.61 - 0.96) | 0.001 |
| L98 : Other disorders of skin and subcutaneous tissue, not elsewhere classified | 1.38 (0.74 - 2.57) | 8.414 | 0.75 (0.62 - 0.91) | <0.0001 | 1.45 (0.73 - 2.90) | 6.721 | 0.7 (0.57 - 0.86) | <0.0001 |
| M10 : Gout | 1.11 (0.45 - 2.73) | 113.835 | 0.53 (0.34 - 0.81) | <0.0001 | * | * | 0.53 (0.33 - 0.85) | <0.0001 |
| M13 : Other arthritis | 1.26 (0.65 - 2.43) | 31.588 | 0.75 (0.52 - 1.10) | 0.768 | 1.11 (0.52 - 2.33) | 96.314 | 0.69 (0.45 - 1.04) | 0.106 |
| M19 : Other and unspecified osteoarthritis | 1.2 (0.86 - 1.67) | 6.208 | 0.73 (0.58 - 0.92) | <0.0001 | 1.02 (0.70 - 1.48) | 130.618 | 0.68 (0.52 - 0.87) | <0.0001 |
| M25 : Pain in joint | 1.5 (1.21 - 1.86) | <0.0001 | 0.97 (0.85 - 1.11) | 75.062 | 1.39 (1.10 - 1.75) | <0.0001 | 0.91 (0.79 - 1.05) | 2.355 |
| M47 : Spondylosis | 1.51 (0.97 - 2.34) | 0.067 | 1.2 (0.89 - 1.62) | 4.149 | 1.24 (0.76 - 2.00) | 15.633 | 1.09 (0.78 - 1.52) | 52.915 |
| M51 : Thoracic, thoracolumbar, and lumbosacral intervertebral disc disorders | 1.48 (0.82 - 2.69) | 2.06 | 1.13 (0.77 - 1.66) | 40.575 | 1.23 (0.60 - 2.52) | 42.022 | 1.0 (0.65 - 1.53) | 161.794 |

| | | | | | | | | |
|---|---|---|---|---|---|---|---|---|
| M53 : Cervicobrachial syndrome | 1.85 (1.32 - 2.57) | <0.0001 | 1.15 (0.77 - 1.73) | 33.739 | 1.47 (1.01 - 2.16) | 0.019 | 1.01 (0.65 - 1.57) | 148.958 |
| M54 : Dorsalgia | 1.56 (1.35 - 1.80) | <0.0001 | 1.0 (0.90 - 1.10) | 175.557 | 1.38 (1.18 - 1.61) | <0.0001 | 0.9 (0.81 - 1.01) | 0.057 |
| M62 : Contracture of muscle | 2.09 (1.44 - 3.02) | <0.0001 | 0.98 (0.62 - 1.55) | 161.079 | * | * | * | * |
| M75 : Shoulder lesions | 1.28 (0.98 - 1.68) | 0.065 | 0.89 (0.70 - 1.12) | 9.592 | 1.11 (0.82 - 1.51) | 31.232 | 0.78 (0.60 - 1.03) | 0.106 |
| M77 : Other enthesopathies | 1.38 (1.07 - 1.77) | 0.0003 | 0.77 (0.62 - 0.96) | 0.0009 | 1.11 (0.84 - 1.48) | 23.679 | 0.74 (0.58 - 0.94) | 0.0003 |
| M79 : Pain in limb, foot / Myalgia | 1.72 (1.40 - 2.11) | <0.0001 | 1.1 (1.00 - 1.22) | 0.057 | 1.46 (1.16 - 1.83) | <0.0001 | 0.96 (0.86 - 1.07) | 24.494 |
| M81 : Osteoporosis without current pathological fracture | 1.35 (0.73 - 2.48) | 11.182 | 1.07 (0.74 - 1.53) | 90.825 | 1.21 (0.61 - 2.39) | 47.598 | 1.0 (0.67 - 1.49) | 160.351 |
| N30 : Cystitis | 1.68 (1.34 - 2.12) | <0.0001 | 1.22 (1.07 - 1.39) | <0.0001 | 1.5 (1.16 - 1.94) | <0.0001 | 1.21 (1.05 - 1.39) | 0.0001 |
| N39 : Urinary tract infection, site not specified | 1.79 (1.27 - 2.51) | <0.0001 | 1.47 (1.27 - 1.70) | <0.0001 | 1.68 (1.14 - 2.46) | <0.0001 | 1.38 (1.18 - 1.61) | <0.0001 |
| N64 : Other disorders of breast | 2.15 (1.24 - 3.74) | <0.0001 | 0.88 (0.70 - 1.11) | 6.903 | 1.48 (0.78 - 2.79) | 3.189 | 0.77 (0.60 - 1.00) | 0.033 |
| N76 : Other inflammation of vagina and vulva | 1.57 (0.92 - 2.69) | 0.233 | 0.86 (0.61 - 1.21) | 16.12 | 1.38 (0.75 - 2.53) | 7.428 | 0.83 (0.58 - 1.19) | 7.908 |
| N91 : Absent, scanty and rare menstruation | 1.62 (0.99 - 2.63) | 0.03 | 0.85 (0.65 - 1.10) | 3.005 | 1.22 (0.70 - 2.13) | 28.108 | 0.87 (0.66 - 1.15) | 8.932 |
| N92 : Excessive, frequent and irregular menstruation | 1.7 (1.10 - 2.62) | 0.0005 | 0.85 (0.73 - 0.98) | 0.002 | 1.45 (0.91 - 2.33) | 0.447 | 0.86 (0.73 - 1.00) | 0.018 |
| N94 : Pain and other conditions associated with female genital | 1.54 (0.96 - 2.48) | 0.084 | 0.99 (0.78 - 1.27) | 175.205 | 1.56 (0.95 - 2.56) | 0.113 | 0.99 (0.77 - 1.27) | 146.078 |

| | | | | | | | | |
|---|---|---|---|---|---|---|---|---|
| organs and menstrual cycle | | | | | | | | |
| N95 : Menopausal and other perimenopausal disorders | 1.18 (0.82 - 1.70) | 14.332 | 0.92 (0.77 - 1.09) | 9.577 | 1.0 (0.66 - 1.51) | 156.467 | 0.88 (0.73 - 1.07) | 2.065 |
| R05 : Cough | 1.24 (1.04 - 1.47) | 0.0005 | 1.04 (0.71 - 1.55) | 126.173 | 1.2 (1.01 - 1.44) | 0.014 | 0.96 (0.62 - 1.47) | 113.26 |
| R06 : Abnormalities of breathing | 1.39 (0.90 - 2.15) | 0.61 | 0.79 (0.56 - 1.11) | 1.435 | 1.33 (0.82 - 2.17) | 3.889 | 0.7 (0.48 - 1.02) | 0.061 |
| R07 : Pain in throat and chest | 1.32 (0.97 - 1.81) | 0.115 | 0.82 (0.63 - 1.08) | 1.251 | 1.2 (0.84 - 1.70) | 9.06 | 0.71 (0.52 - 0.96) | 0.003 |
| R10 : Abdominal and pelvic pain | 1.28 (1.06 - 1.55) | 0.0001 | 0.88 (0.74 - 1.05) | 1.303 | 1.18 (0.97 - 1.45) | 0.285 | 0.84 (0.70 - 1.01) | 0.077 |
| R20 : Paresthesia of skin | 14.81 (9.74 - 22.52) | <0.0001 | 9.45 (7.42 - 12.04) | <0.0001 | * | * | * | * |
| R21 : Rash and other nonspecific skin eruption | 1.4 (0.80 - 2.45) | 3.89 | 1.04 (0.82 - 1.31) | 103.362 | 1.02 (0.51 - 2.05) | 143.795 | 0.93 (0.72 - 1.20) | 46.048 |
| R25 : Abnormal involuntary movements | 1.47 (0.89 - 2.45) | 0.654 | 2.32 (1.80 - 2.99) | <0.0001 | 1.21 (0.66 - 2.19) | 37.357 | * | * |
| R42 : Dizziness and giddiness | 2.57 (1.93 - 3.41) | <0.0001 | 2.08 (1.78 - 2.41) | <0.0001 | * | * | * | * |
| R51 : Headache | 1.48 (1.18 - 1.87) | <0.0001 | 1.28 (1.01 - 1.62) | 0.019 | 1.17 (0.90 - 1.51) | 3.766 | 1.14 (0.88 - 1.48) | 8.734 |
| R52 : Pain, unspecified | 1.66 (1.26 - 2.18) | <0.0001 | 0.96 (0.67 - 1.39) | 132.714 | 1.32 (0.96 - 1.82) | 0.172 | 0.83 (0.55 - 1.25) | 13.569 |
| R53 : Malaise and fatigue | 1.76 (1.47 - 2.10) | <0.0001 | 1.45 (1.13 - 1.87) | <0.0001 | 1.55 (1.28 - 1.88) | <0.0001 | 1.21 (0.92 - 1.59) | 1.442 |
| R55 : Syncope and collapse | 2.01 (1.13 - 3.58) | 0.0007 | 1.07 (0.74 - 1.55) | 96.066 | 1.33 (0.70 - 2.54) | 15.262 | 0.93 (0.62 - 1.41) | 82.984 |
| R59 : Enlarged lymph nodes | 1.76 (1.12 - 2.78) | 0.0004 | 0.86 (0.54 - 1.39) | 44.605 | 1.68 (1.04 - 2.69) | 0.006 | * | * |

| | | | | | | | | |
|---|---|---|---|---|---|---|---|---|
| R60 : Edema, not elsewhere classified | 1.47 (0.88 - 2.47) | 0.702 | 0.99 (0.68 - 1.44) | 171.227 | 1.36 (0.75 - 2.44) | 8.025 | 0.79 (0.51 - 1.22) | 6.796 |
| R63 : Abnormal weight loss or gain | 1.7 (1.04 - 2.78) | 0.007 | 1.03 (0.67 - 1.59) | 151.349 | 1.55 (0.88 - 2.70) | 0.502 | 0.81 (0.49 - 1.33) | 17.177 |
| S83 : Dislocation and sprain of joints and ligaments of knee | 1.51 (0.81 - 2.80) | 2.156 | 0.7 (0.44 - 1.12) | 0.717 | 1.27 (0.63 - 2.54) | 31.385 | * | * |
| S93 : Dislocation and sprain of joints and ligaments at ankle, foot and toe level | 1.69 (1.17 - 2.45) | <0.0001 | 1.06 (0.78 - 1.45) | 86.342 | 1.41 (0.94 - 2.10) | 0.204 | 0.97 (0.69 - 1.36) | 115.051 |
| T14 : Injury of unspecified body region | 1.66 (1.34 - 2.05) | <0.0001 | 0.93 (0.76 - 1.14) | 36.223 | 1.34 (1.06 - 1.69) | 0.0003 | 0.87 (0.70 - 1.08) | 2.442 |
| T78 : Adverse effects, not elsewhere classified | 1.28 (1.02 - 1.61) | 0.005 | 0.92 (0.68 - 1.25) | 57.986 | 1.24 (0.97 - 1.57) | 0.149 | 0.92 (0.67 - 1.28) | 57.517 |
| Z01 : Encounter for other special examination without complaint, suspected or reported diagnosis | 1.16 (0.71 - 1.91) | 44.452 | 1.11 (0.70 - 1.74) | 73.733 | 0.75 (0.39 - 1.44) | 15.08 | * | * |
| Z02 : Encounter for administrative examination | 1.37 (1.13 - 1.65) | <0.0001 | 1.24 (1.04 - 1.48) | 0.0009 | 1.2 (0.98 - 1.48) | 0.106 | 1.03 (0.84 - 1.27) | 87.774 |
| Z13 : Encounter for screening for other diseases and disorders | 1.04 (0.66 - 1.65) | 128.032 | 0.65 (0.41 - 1.05) | 0.128 | 1.04 (0.64 - 1.70) | 120.961 | * | * |
| Z30 : Encounter for contraceptive management | 1.35 (1.08 - 1.68) | <0.0001 | 0.98 (0.89 - 1.09) | 107.646 | 1.4 (1.11 - 1.76) | <0.0001 | 1.06 (0.96 - 1.18) | 5.031 |
| Z76 : Persons encountering health services in | 1.22 (0.99 - 1.50) | 0.065 | 0.81 (0.59 - 1.11) | 1.813 | 1.12 (0.89 - 1.40) | 11.386 | 0.84 (0.59 - 1.20) | 10.692 |

other
circumstances

| | Primary analysis | | | | | | Sensitivity analysis | | | | | |
|---|---|---|---|---|---|---|---|---|---|---|---|---|
| | FR | | | UK | | | FR | | | UK | | |
| | No. (%) [MS:controls] | OR (Adjusted 95% CI) | Adjusted p value | No. (%) [MS:controls] | OR (Adjusted 95% CI) | Adjusted p value | No. (%) [MS:controls] | OR (Adjusted 95% CI) | Adjusted p value | No. (%) [MS:controls] | OR (Adjusted 95% CI) | Adjusted p value |
| F32 : Major depressive disorder, single episode | 789 (18.1): 1350 (10.8) | 1.84 (1.52 - 2.21) | <0.0001 | 3801 (24.0): 8297 (19.6) | 1.24 (1.14 - 1.36) | <0.0001 | 630 (14.4): 1139 (9.1) | 1.62 (1.32 - 1.98) | <0.0001 | 3295 (20.8): 7030 (16.6) | 1.22 (1.11 - 1.34) | <0.0001 |
| F52 : Sexual dysfunction | 154 (3.6): 246 (2.0) | 1.92 (1.29 - 2.86) | <0.0001 | 388 (2.5): 650 (1.5) | 1.54 (1.19 - 1.99) | <0.0001 | 123 (2.8): 209 (1.7) | 1.69 (1.10 - 2.62) | 0.0008 | 305 (1.9): 515 (1.2) | 1.47 (1.11 - 1.95) | <0.0001 |
| G40 : Epilepsy and recurrent seizures | 61 (1.4): 71 (0.6) | 2.54 (1.30 - 4.97) | <0.0001 | 188 (1.2): 363 (0.9) | 1.43 (1.00 - 2.04) | 0.021 | ..* | ..* | ..* | 167 (1.1): 308 (0.7) | 1.46 (1.00 - 2.12) | 0.022 |
| H81 : Disorders of vestibular function | 144 (3.3): 184 (1.5) | 2.22 (1.44 - 3.43) | <0.0001 | 330 (2.1): 557 (1.3) | 1.49 (1.13 - 1.97) | <0.0001 | .* | .* | .* | .* | .* | .* |
| K59 : Functional intestinal disorders | 406 (9.3): 655 (5.2) | 1.9 (1.47 - 2.44) | <0.0001 | 1171 (7.4): 1845 (4.4) | 1.58 (1.35 - 1.84) | <0.0001 | 340 (7.8): 591 (4.7) | 1.66 (1.27 - 2.17) | <0.0001 | 904 (5.7): 1427 (3.4) | 1.5 (1.27 - 1.78) | <0.0001 |
| N30 : Cystitis | 478 (11.0): 855 (6.8) | 1.68 (1.34 - 2.12) | <0.0001 | 1422 (9.0): 2951 (7.0) | 1.22 (1.07 - 1.39) | <0.0001 | 367 (8.4): 687 (5.5) | 1.5 (1.16 - 1.94) | <0.0001 | 1176 (7.4): 2385 (5.6) | 1.21 (1.05 - 1.39) | 0.0001 |
| N39 : Urinary tract infection, site not specified | 217 (5.0): 348 (2.8) | 1.79 (1.27 - 2.51) | <0.0001 | 1218 (7.7): 2263 (5.4) | 1.47 (1.27 - 1.70) | <0.0001 | 165 (3.8): 268 (2.1) | 1.68 (1.14 - 2.46) | <0.0001 | 997 (6.3): 1904 (4.5) | 1.38 (1.18 - 1.61) | <0.0001 |
| R20 : Paresthesia of skin | 512 (11.7): 103 (0.8) | 14.81 (9.74 - 22.52) | <0.0001 | 1220 (7.7): 363 (0.9) | 9.45 (7.42 - 12.04) | <0.0001 | .* | .* | .* | .* | .* | .* |
| R42 : Dizziness and giddiness | 372 (8.5): 415 (3.3) | 2.57 (1.93 - 3.41) | <0.0001 | 1300 (8.2): 1667 (4.0) | 2.08 (1.78 - 2.41) | <0.0001 | .* | .* | .* | .* | .* | .* |
| R51 : Headache | 464 (10.6): 908 (7.2) | 1.48 (1.18 - 1.87) | <0.0001 | 403 (2.6): 844 (2.0) | 1.28 (1.00 - 1.62) | 0.019 | 333 (7.6): 776 (6.2) | 1.17 (0.90 - 1.51) | 1 | 324 (2.0): 730 (1.7) | 1.14 (0.88 - 1.48) | 1 |
| R53 : Malaise and fatigue | 891 (20.4): 1574 (12.5) | 1.76 (1.47 - 2.10) | <0.0001 | 376 (2.4): 692 (1.6) | 1.45 (1.13 - 1.87) | <0.0001 | 703 (16.1): 1301 (10.4) | 1.55 (1.28 - 1.88) | <0.0001 | 294 (1.9): 633 (1.5) | 1.21 (0.92 - 1.59) | 1 |

*Table 6 ORs for all health problems individually associated with a future diagnosis of MS for the five-year period leading up to the index date. In the primary analysis, we used the first recorded demyelinating event as the index date, whereas, in the sensitivity analysis, the first symptom suggestive of MS was used as the index date. .* cannot be calculated because the health condition is used to define the index date in the sensitivity analysis. ..* cannot be calculated because too few presentations were recorded.*

| | | Sex | | Age at onset | | | Year of diagnosis | | |
|---|---|---|---|---|---|---|---|---|---|
| | Whole cohort | Women | Men | <40 | [40,53] | >53 | <2010 | [2010,2017] | >2017 |
| **Before diagnosis** | | | | | | | | | |
| F32 : Major depressive disorder, single episode | 1.62 (1.32 - 1.98)** | 1.44 (1.22 - 1.71)** | 2.13 (1.54 - 2.93)** | 1.93 (1.46 - 2.55)** | 1.54 (1.21 - 1.96)** | 1.32 (1.01 - 1.73)* | 1.56 (1.26 - 1.93)** | 1.58 (1.23 - 2.03)** | 1.45 (0.95 - 2.22) |
| F52 : Sexual dysfunction | 1.69 (1.10 - 2.62)* | 1.7 (1.14 - 2.55)* | 1.47 (0.86 - 2.49) | 1.76 (0.92 - 3.38) | 1.72 (1.06 - 2.78)* | 1.29 (0.71 - 2.32) | 1.42 (0.92 - 2.18) | 1.94 (1.13 - 3.32)* | 1.71 (0.48 - 6.13) |
| K59 : Functional intestinal disorders | 1.66 (1.27 - 2.17)** | 1.52 (1.21 - 1.90)** | 2.04 (1.35 - 3.09)** | 1.6 (1.15 - 2.23)* | 1.9 (1.34 - 2.69)** | 1.41 (0.99 - 2.00)* | 1.48 (1.12 - 1.95)* | 1.92 (1.35 - 2.72)** | 1.44 (0.84 - 2.45) |
| N30 : Cystitis | 1.5 (1.16 - 1.94)** | 1.32 (1.08 - 1.62)* | 3.25 (1.75 - 6.04)** | 1.38 (1.00 - 1.89)* | 1.46 (1.05 - 2.03)* | 1.45 (1.03 - 2.03)* | 1.58 (1.19 - 2.10)** | 1.29 (0.94 - 1.77) | 1.24 (0.79 - 1.94) |
| N39 : Urinary tract infection, site not specified | 1.68 (1.14 - 2.46)** | 1.55 (1.14 - 2.10)* | 2.21 (1.02 - 4.78)* | 1.75 (1.07 - 2.85)* | 1.27 (0.76 - 2.15) | 1.83 (1.15 - 2.93)* | 1.8 (1.14 - 2.85)* | 1.43 (0.92 - 2.21) | 1.71 (0.88 - 3.36) |
| **After diagnosis** | | | | | | | | | |
| F32 : Major depressive disorder, single episode | 1.34 (1.16 - 1.54)** | 1.3 (1.10 - 1.53)** | 1.45 (1.06 - 1.98)* | 1.67 (1.28 - 2.19)** | 1.32 (1.06 - 1.64)* | 1.1 (0.83 - 1.47) | 1.41 (1.17 - 1.70)** | 1.3 (1.01 - 1.67)* | 0.97 (0.51 - 1.84) |
| F52 : Sexual dysfunction | 1.41 (1.01 - 2.01)* | 1.28 (0.73 - 2.24) | 1.52 (0.95 - 2.44) | 1.63 (0.76 - 3.50) | 1.44 (0.85 - 2.45) | 1.3 (0.68 - 2.49) | 1.43 (0.95 - 2.15) | 1.34 (0.59 - 3.00) | 5.11 (0.25 - 106.07) |
| K59 : Other functional intestinal disorders | 1.98 (1.63 - 2.41)** | 1.77 (1.41 - 2.22)** | 2.78 (1.86 - 4.16)** | 1.85 (1.25 - 2.74)** | 2.35 (1.68 - 3.28)** | 1.91 (1.39 - 2.61)** | 2.02 (1.57 - 2.59)** | 2.02 (1.42 - 2.86)** | 2.06 (0.75 - 5.66) |
| N30 : Cystitis | 1.56 (1.31 - 1.86)** | 1.42 (1.19 - 1.71)** | 6.55 (2.62 - 16.35)** | 1.5 (1.09 - 2.05)* | 1.55 (1.16 - 2.06)** | 1.77 (1.28 - 2.45)** | 1.65 (1.32 - 2.07)** | 1.33 (0.97 - 1.82) | 2.0 (0.88 - 4.53) |
| N39 : Other disorders of urinary system | 2.21 (1.74 - 2.82)** | 2.11 (1.63 - 2.74)** | 2.94 (1.52 - 5.68)** | 2.24 (1.40 - 3.58)** | 2.29 (1.55 - 3.40)** | 2.25 (1.50 - 3.38)** | 2.39 (1.74 - 3.28)** | 1.9 (1.27 - 2.83)** | 2.85 (0.84 - 9.66) |

*Table 7 ORs for all health troubles individually associated with MS in the sensitivity analysis cohort at 5 years before and after the first symptoms in the French cohort. We used the whole sensitivity cohort, sex cohorts, age at onset cohorts, and*

*year at diagnosis cohorts. Results are shown for the French cohort. .* cannot be calculated because an insufficient number of presentations was recorded. * for ORs associated to corrected p-values between 0.05 and 0.0001. ** for ORs associated to corrected p-values below 0.0001.*

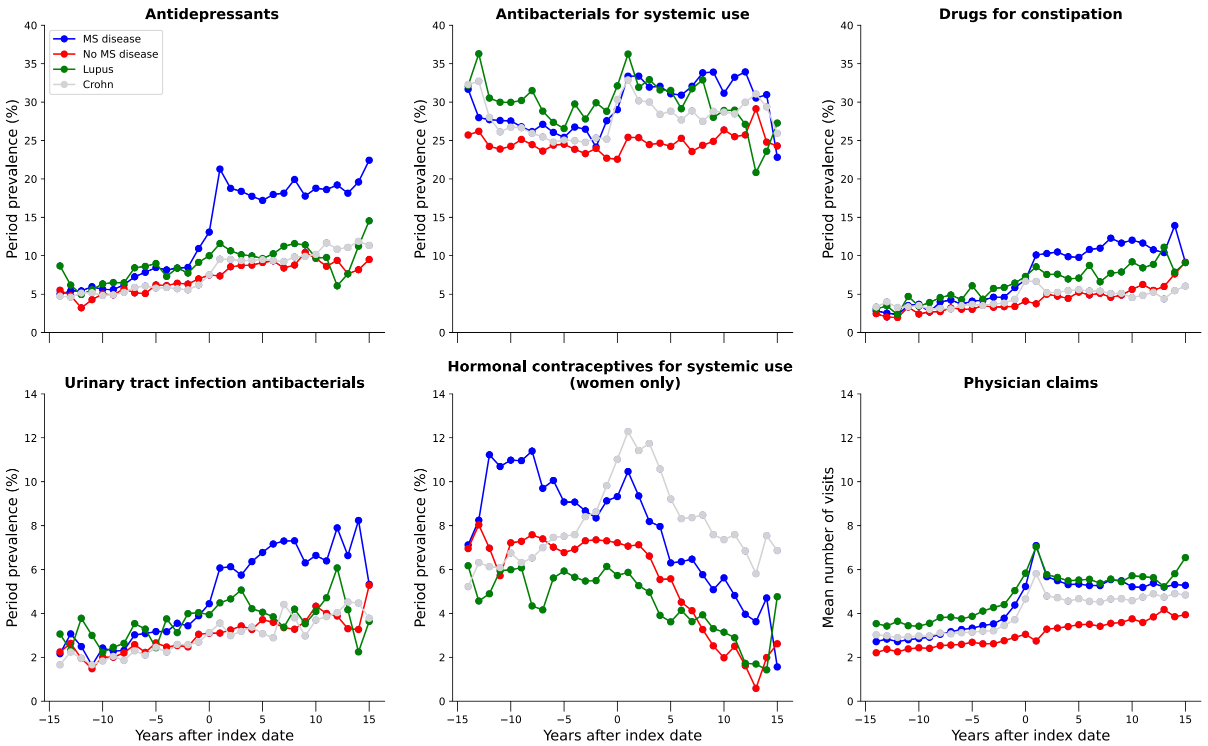


*Figure 3 Period prevalence (%) of prescriptions associated to depression, urinary tract infections, bacterial infections, constipation, and hormonal contraceptives. In the bottom left, the mean number of visits per year is displayed.*
